# Randomized, Double-Blind, Placebo-Controlled Trial of MUC1 Peptide Vaccine for Prevention of Recurrent Colorectal Adenoma

**DOI:** 10.1101/2022.10.05.22280474

**Authors:** Robert E. Schoen, Lisa A. Boardman, Marcia Cruz-Correa, Ajay Bansal, David Kastenberg, Chin Hur, Lynda Dzubinski, Sharon F. Kaufman, Luz M. Rodriguez, Ellen Richmond, Asad Umar, Eva Szabo, Andres Salazar, John McKolanis, Pamela Beatty, Reetesh K. Pai, Aatur D. Singhi, Camille M. Jacqueline, Riuye Bao, Brenda Diergaarde, Ryan P. McMurray, Carrie Strand, Nathan R. Foster, David M. Zahrieh, Paul J. Limburg, Olivera J. Finn

## Abstract

**Objective:** Vaccines against antigens expressed on adenomas could prevent new adenoma formation. We assessed whether a MUC1 peptide vaccine produces an immune response and prevents subsequent colonic adenoma formation.

**Design:** Multicenter, double blind, placebo-controlled randomized trial in individuals age 40-70 with diagnosis of an advanced adenoma ≤1 year from randomization. Vaccine was administered at 0, 2, and 10 weeks with a booster injection at week 53. Adenoma recurrence was assessed ≥1 year from randomization. The primary endpoint was vaccine immunogenicity at 12 weeks defined by anti-MUC1 ratio ≥2.0.

**Results:** 53 participants received the MUC1 vaccine and 50 placebo. 13/52 (25%) of MUC1 vaccine recipients had a ≥2-fold increase in MUC1 IgG (range 2.9-17.3) at week 12 vs. 0/50 placebo recipients (1-sided Fisher’s exact P<0.0001). Of the 13 responders at week 12, 11 (84.6%) had a ≥2-fold increase in MUC1 IgG with the booster and were considered immune responders. A recurrent adenoma was observed in 31 of 47 (66.0%) in the placebo group vs. 27 of 48 (56.3%) participants in the MUC1 group (adjusted relative risk (aRR) = 0.83 [95% CI, 0.60-1.14], P=0.25). Adenoma recurrence occurred in 3/11 (27.3%) immune responders, (aRR = 0.41 [95% CI, 0.15-1.11], P=0.08). Vaccine recipients had more injection site reactions than placebo recipients, but there was no difference in serious adverse events.

**Conclusion:** An immune response was observed only in vaccine recipients. Overall adenoma recurrence was not different than placebo, but a 38% absolute reduction in adenoma recurrence was observed in immune responders.

ClinicalTrials.gov Identifier: NCT02134925.

https://clinicaltrials.gov/ct2/show/NCT02134925

**Key Points:** *What is already known:* Antigens expressed on colonic adenomas are potential targets for immunopreventive vaccines. An effective vaccine could prevent subsequent adenoma formation.

*What this Study Adds:* In this multicenter, double blind, placebo-controlled randomized trial, MUC1 vaccine recipients developed an immune response. Overall adenoma recurrence was not different than placebo, but a 38% absolute reduction in adenoma recurrence was observed in immune responders.

*How this study might affect research, practice or policy:* Vaccine immunoprevention is a potential new frontier to colorectal cancer prevention.

## Introduction

Endoscopic removal of adenomatous polyps, the precursor lesion of colorectal cancer, reduces subsequent colorectal cancer (CRC) incidence (1). However, removing all adenomas is an inefficient means of preventing cancer, because there are many more adenomas than cancers, and most adenomas will not evolve to malignancy. Moreover, adenomatous polyp recurrence rates are high, and repeated colonoscopic surveillance to monitor and remove recurrent adenomas is expensive, invasive, and associated with medical risk. Methods that would either prevent adenomas from forming or prevent them from evolving into malignancy would be welcome.

Clinical trials of chemoprevention to prevent recurrence of adenomatous polyps with agents such as aspirin (2), calcium (3), folate (4), DMFO (5), dietary manipulation (6), have been performed. Most have demonstrated limited or no benefit. The most promising agents, including aspirin or DFMO, are limited by side effects and by the burden of compliance, as chemoprevention agents must be taken regularly.

Immunoprevention with vaccines, to target and eliminate pre-malignant precursors, is a potentially safe and effective approach to cancer control (7). Moreover, because of the specificity of the immune response and its long-term memory, immunoprevention offers the potential for prolonged protection. Targeting antigens aberrantly expressed on cancers and their precursors offers the potential for a relatively non-invasive and non-toxic risk-reduction strategy.

MUC1 mucin is a high molecular weight transmembrane glycoprotein expressed in normal epithelial cells, polarized to the apical surface, and extensively glycosylated (8). Cancer patients with MUC1 positive tumors produce MUC1-specific antibodies and T cells at low levels (9, 10). MUC1-based vaccines raise the immune response to therapeutic levels (11, 12). However, immunosuppressive forces in the tumor microenvironment blunt the immune response (13-15), suggesting that vaccines administered in the pre-malignant phase when immunosuppression is not expected, might generate a more robust immune response.

Individuals with a history of advanced adenomatous polyps are at a 3-fold increased risk of CRC compared to those without adenomas (16). Colonic adenomas, like CRC, express the abnormal form of MUC1 (17). In a pilot study, MUC1 vaccine in participants with a history of advanced adenomas resulted in a 2- to 40-fold increase in anti-MUC1 IgG in 44% (17/39), with a robust 1 year memory response, and without significant toxicity (11). We performed a trial of MUC1 vaccine versus placebo in individuals with advanced adenomas within 1 year of removal. The objective was to evaluate the immune response to a MUC1 vaccine and its effect on recurrence of adenomatous polyps.

## Methods

### Study Design

This was a randomized, double-blind, placebo controlled, multicenter trial. Eligible participants with ≥1 advanced adenoma within 1 year were randomly assigned in a 1:1 ratio to receive vaccine or placebo. Participants were enrolled at six clinical centers, the Mayo Clinic, Rochester MN; the University of Pittsburgh Medical Center, Pittsburgh PA; the University of Puerto Rico, San Juan PR; the Veterans Administration, Kansas City, KS; Thomas Jefferson University, Philadelphia PA; and Massachusetts General Hospital, Boston MA. The trial was administratively coordinated by the National Cancer Institute Division of Cancer Prevention through the Cancer Prevention Network at the Mayo Clinic. All authors had access to the study data and reviewed and approved the final manuscript. Patients or the public were not involved in the design, conduct, reporting, or dissemination plans of this research.

### Inclusion Criteria

Participants were 40 - 70 years of age at the time of randomization, had no prior history of colorectal cancer, no history of heritable colorectal cancer syndromes, no history of malignancy within the previous 5 years other than non-melanoma skin cancer, no nonalcoholic steatohepatitis (NASH) with a nonalcoholic fatty liver disease (NAFLD) activity score ≥ 5, no history of auto-immune disease nor current or planned use of immunomodulators, and no corticosteroid use within the previous 12 weeks. All participants had an advanced adenoma (AA) within 1 year prior to randomization, defined as: ≥ 1 cm in size, with villous or tubulovillous histology, or with severe or high-grade dysplasia. Complete removal of all adenomatous lesions and normal blood testing within defined parameters for hematologic, renal and liver function, and anti-nuclear antibody was required.

### MUC1 Vaccine

The MUC1 vaccine consisted of a 100-amino acid synthetic MUC1 (18) (Methods supplement M1) admixed with an adjuvant, toll-like receptor 3 (TLR3) agonist, polyinosinic-polycytidylic acid (Poly ICLC - Hiltonol®), supplied by Oncovir Inc. (Washington, DC). Poly ICLC is a synthetic, non-replicating double-stranded ribonucleic acid (dsRNA), with no specific genetic message. It acts as a viral mimic with broad innate and adaptive immune enhancing, adjuvant, antiviral and antiproliferative effects (19, 20).

Vaccine or placebo was administered subcutaneously, blinded to content, in the upper thigh. Vaccine or placebo was administered at week 0, 2 and 10 with a booster dose administered at week 53. Blood was drawn just prior to vaccination at week 0, 2, 10, and 52, at week 12 and 55 for assessment of immune response, and within ± 4 weeks of the colonoscopy to evaluate for adenoma recurrence.

### Immune and Colonoscopy Endpoints

An anti-MUC1 IgG response requires activation of MUC1-specific helper T cells to promote isotype switching from IgM to IgG, hence indirectly measures T cell immunity. The primary endpoint was MUC1 IgG levels at week 12 in vaccine vs. placebo recipients. An anti-MUC1 IgG ratio of ≥2.0 at 12 weeks (week 12/week 0) was defined as an immune response. Anti-MUC1 IgG levels at week 55 (T55/T52) were also measured after booster injection at week 53. Individuals having an anti-MUC1 IgG ratio of ≥2.0 at 12 weeks and at 55 weeks were considered immune responders.

Adenoma recurrence at the first colonoscopy >1 year post initial vaccination was the primary clinical outcome. The standard for individuals with advanced adenoma is to undergo surveillance colonoscopy at 3 years, but follow-up colonoscopy timing was determined by the treating endoscopist.

Pre-specified, alternative clinical endpoints including adenoma outcome in immune responders, excluding recurrent advanced adenomas that were in the same segment as the baseline advanced adenoma which could represent a residual as opposed to a new recurrence, and restricting recurrence to adenomas >5mm since diminutive adenomas can be missed (21).

### Adverse Events (AEs)

The NCI common terminology criteria for adverse events (CTCAE) version 4.0 was used to monitor toxicity.

### Measurement of Anti-MUC1 IgG, Myeloid-Derived Suppressor Cells (MDSC), and Cytokines

Enzyme-linked Immunosorbent Assay (ELISA) was used to measure plasma anti-MUC1 IgG levels (11). MDSC subpopulations were characterized based on their cell surface markers into polymorphonuclear (PMN)-MDSC, monocytic (M)-MDSCand early (e)-MDSC (22). Bead-based multiplex cytokine assays were used to measure IL-1*β*, INF-*α*, IL-6, INF-*γ*, MCP1, TNF-*α*, IL-8, IL-10, IL-12, IL-17, IL-18, IL-23, IL-33. The cytokines in this panel were chosen based on their previously reported roles in inflammation and cancer (23, 24). IL-1*β*, INF-*α*, IL-6, INF-*γ*, MCP1, TNF*α* and IL-8 (or CXCL8) are proinflammatory cytokines that are known to suppress immunity and IL-10 can suppress both innate cell-mediated and adaptive immunity. IL-12, IL-17, IL-18 and IL-23 promote type 1 immunity while IL-33 promotes type 2 immunity (Methods supplement M2).

### Immunohistochemistry (IHC) Staining of Polyps for MUC1 Expression

Formalin-fixed, paraffin-embedded sections were stained using a rabbit monoclonal antibody against MUC1 (1:100 dilution). Stained tissues were evaluated for the extent, intensity, and localization of staining (25) (Methods Supplement M2).

### Immunofluorescence Staining, Image Collection and Analysis

Uniplex immunofluorescence staining was performed manually using the Opal 6-Plex kit (Akoya Biosciences, Marlborough, MA), which uses individual tyramide signal amplification (TSA)-conjugated fluorophores to detect targets. The slides were scanned using the Vectra Polaris spectral imaging system (Akoya Biosciences) (26). A total of 466 regions of interest (ROI) were identified using Phenochart under the supervision of a gastrointestinal-trained pathologist (ADS). Five marker-positive cells were annotated, including CD4+ T cells, CD8+ T cells, CD15+ myeloid cells, CD20+ B cells, and FOXP3+ regulatory T cells (Methods Supplement M2).

### Statistical Analysis

Participants were randomized at the Mayo coordinating center in a 1:1 fashion to MUC1 or placebo using the Pocock-Simon dynamic allocation procedure (27), which balances the marginal distributions of the stratification (28). Stratification factors included number of adenomatous polyps removed (>=3 vs. < 3), gender, and treatment site.

The primary endpoint was to compare the ratio of the week 12 to week 0 IgG levels between MUC1 vaccine and placebo. Assuming equal standard deviations (i.e. 10) across the MUC1 and placebo groups, and assuming up to a 10% drop-out rate by year 1, retaining at least 50 evaluable participants per arm yields at least 86% power to detect an increase in the mean IgG ratio from 1 to 6.5 for placebo vs. MUC1 (effect size = 0.55), using a 1-sided t-test with a significance level of 0.05.

Univariate log-binomial models were used to compute estimates of the relative risk (RR). Multivariate logistic regression models were used to compute the adjusted estimates of the odds ratio (OR), which then in turn were used to approximate estimates of the relative risk (RR) by the following formula: 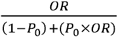, where *P*_0_ indicates incidence of the varying types of adenoma recurrence in the placebo group. Variables included in the multivariate models were body mass index (BMI), gender (male vs. female), nonsteroidal anti-inflammatory drug (NSAID) use (yes vs. no), aspirin use (yes vs. no), multiple baseline adenomas (yes vs. no), and multiple baseline advanced adenomas (yes vs. no). Multivariate associations were assessed by the Wald test. All tests were performed with use of a two-sided alpha level of 0.05, unless otherwise specified. SAS version 9.4 (SAS Institute, Inc.) was used for statistical analysis.

## Results

The Consort diagram for participant flow is shown in Figure 1. A total of 130 participants were pre-registered and signed informed consent. Consented participants underwent additional laboratory testing and evaluation and 110 were confirmed eligible. With both investigator and participant blinded to randomization arm, 7 eligible participants declined to proceed. Of the remaining 103 participants, one withdrew from subsequent study visits at week 2 but remained in the study, leaving 102 (52 in the MUC1 group and 50 in the placebo group) evaluable at week 12. Ninety-five participants (51 in the MUC1 group and 44 in the placebo group) completed testing for booster response/immune memory at 52 and 55 weeks. One participant withdrew after 52 weeks. Ninety-five participants (48 in the MUC1 group and 47 in the placebo group) had an endpoint colonoscopy.

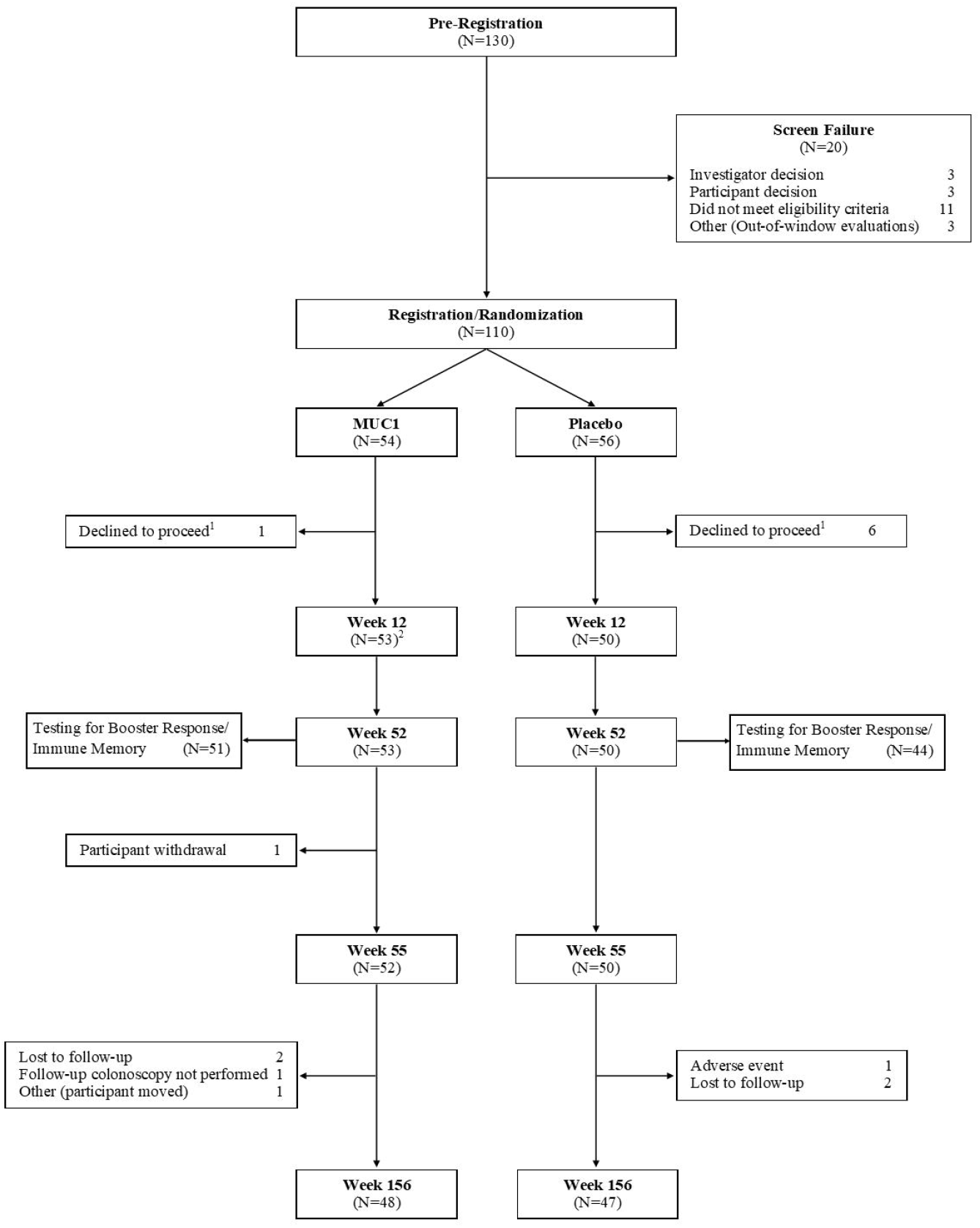

Among the 103 intervention participants, the mean age was 59.4 ± 7.0 years, 62.1% were male, 88.3% were white, and 18.4% were Hispanic, with no significant differences by study arm (Table 1). Clinical characteristics and endoscopic and advanced adenoma findings at the qualifying, baseline colonoscopy were similar between the two study arms (Table 1).

**Table 1.**
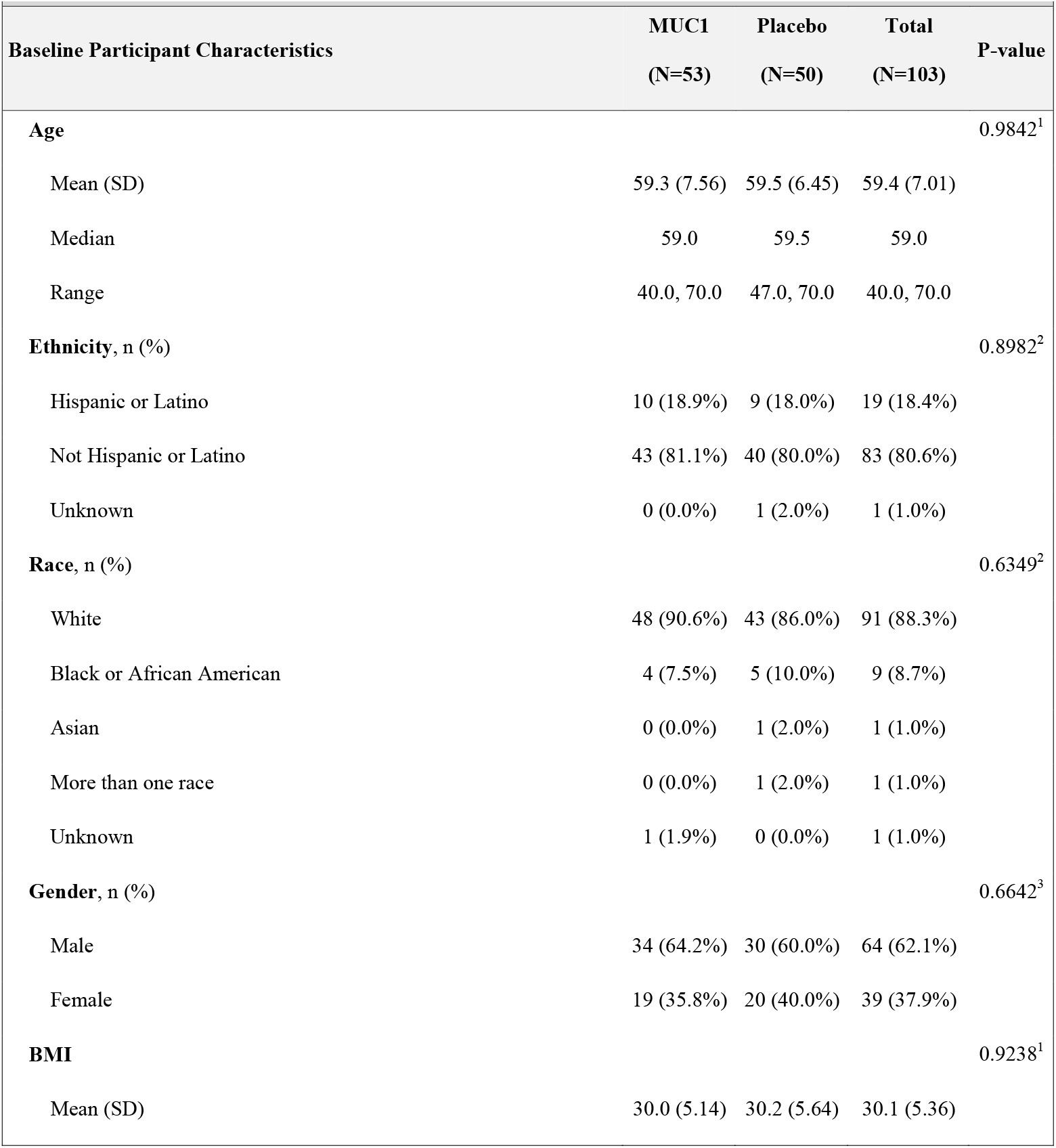
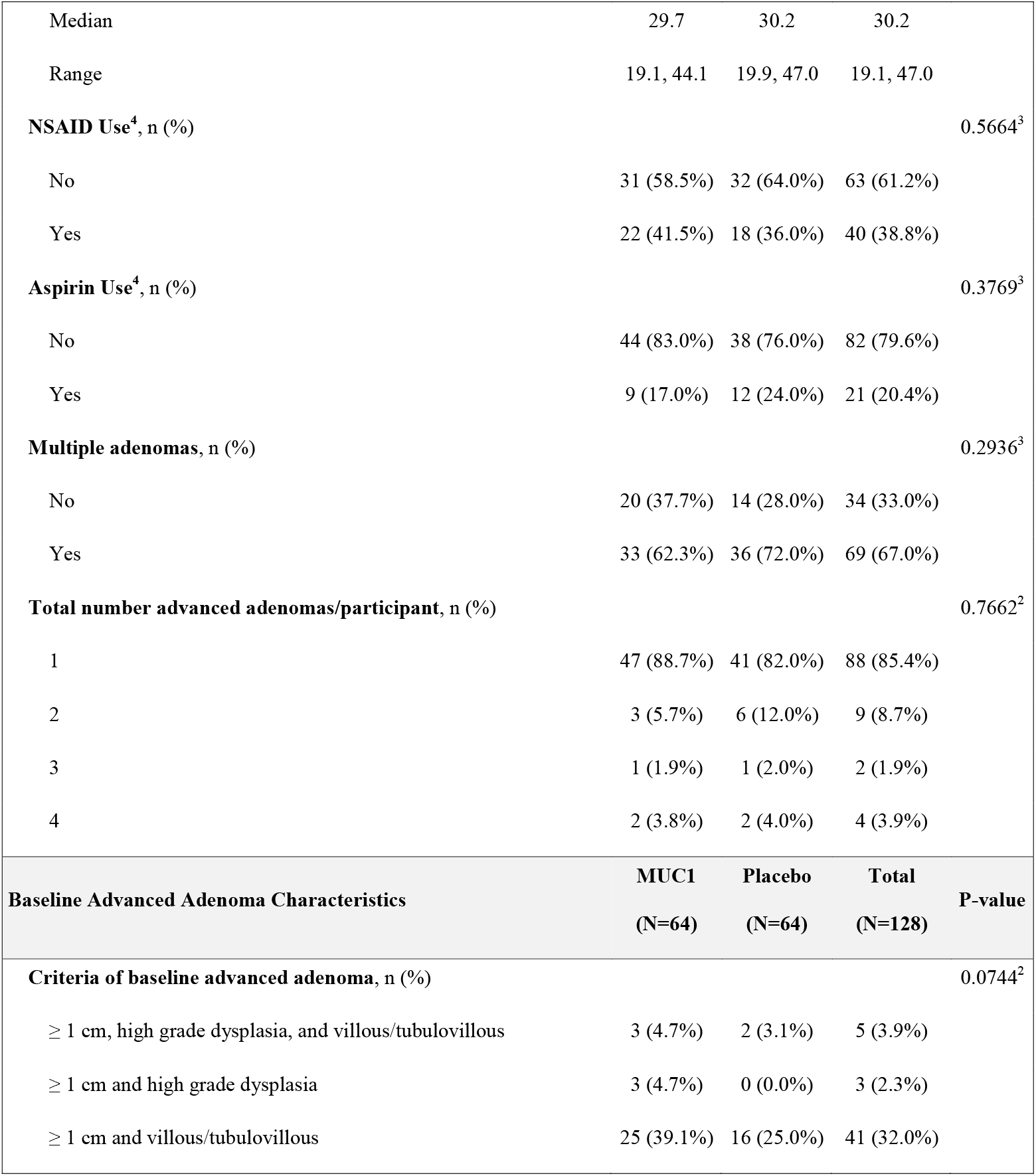
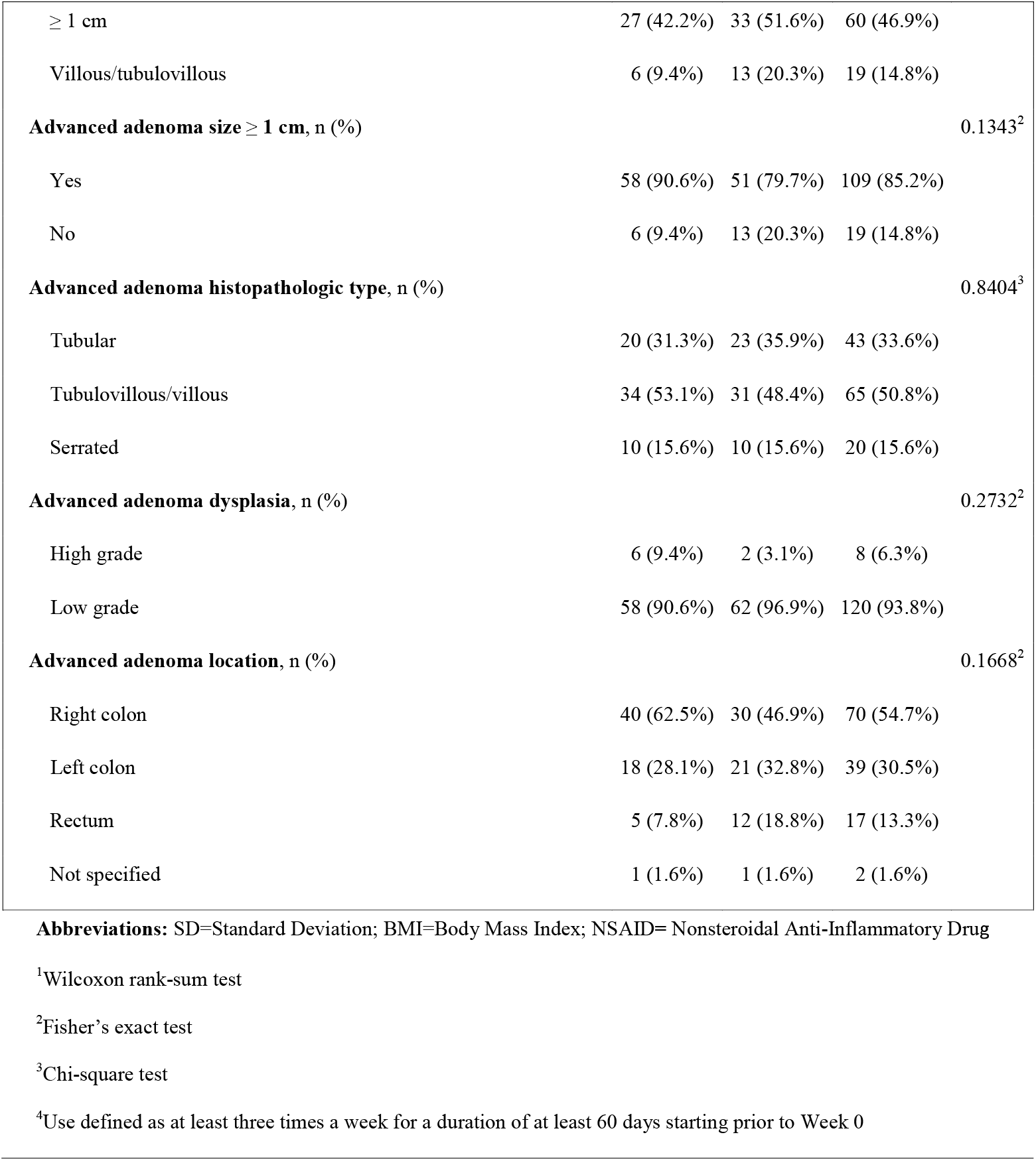
Baseline Participant and Advanced Adenoma Characteristics.

### Vaccine immunogenicity

After injections at 0, 2, and 10 weeks, the mean ± standard deviation (SD) of week 12/week 0 IgG ratio was significantly higher in MUC1 vaccine vs. placebo recipients (3.0 ± 4.31 vs. 1.0 ± 0.19, P=0.0004). An anti-MUC1 IgG ratio of ≥2.0 at 12 weeks was observed in 13/52 (25%) MUC1 vaccine recipients vs. 0/50 placebo recipients (1-sided Fisher’s exact P<0.0001. After the booster injection, an anti-MUC1 IgG ratio of ≥2.0 at week 55/week 52 was observed in 17/51 (33.3%) MUC1 vaccine recipients versus 2/44 (4.5%) in the placebo group (P=0.0003). Of the 13 MUC1 vaccine recipients who had an immune response at week 12, 11 (84.6%) had a ≥2-fold increase in MUC1 IgG at week 55. These 11 participants were classified as immune responders whereas 0/44 participants in the placebo group were immune responders. Figure 2A shows the kinetics of the antibody generation in 13 participants who had an immune response at week 12. Figure 2B shows fold elevation in anti-MUC1 IgG at week 12 (N=13, range 2.9 - 17.3, mean 8.9). Figure 2C shows pre and post vaccination anti-MUC1 IgG levels in the MUC1 vaccine recipients who did not manifest an immune response and figure 2D shows anti-MUC1 IgG levels in the placebo group. Endpoint titers of plasma antibodies at week 12 amongst those with an immune response is shown in Supplemental Figure 1.

**Figure 2:**
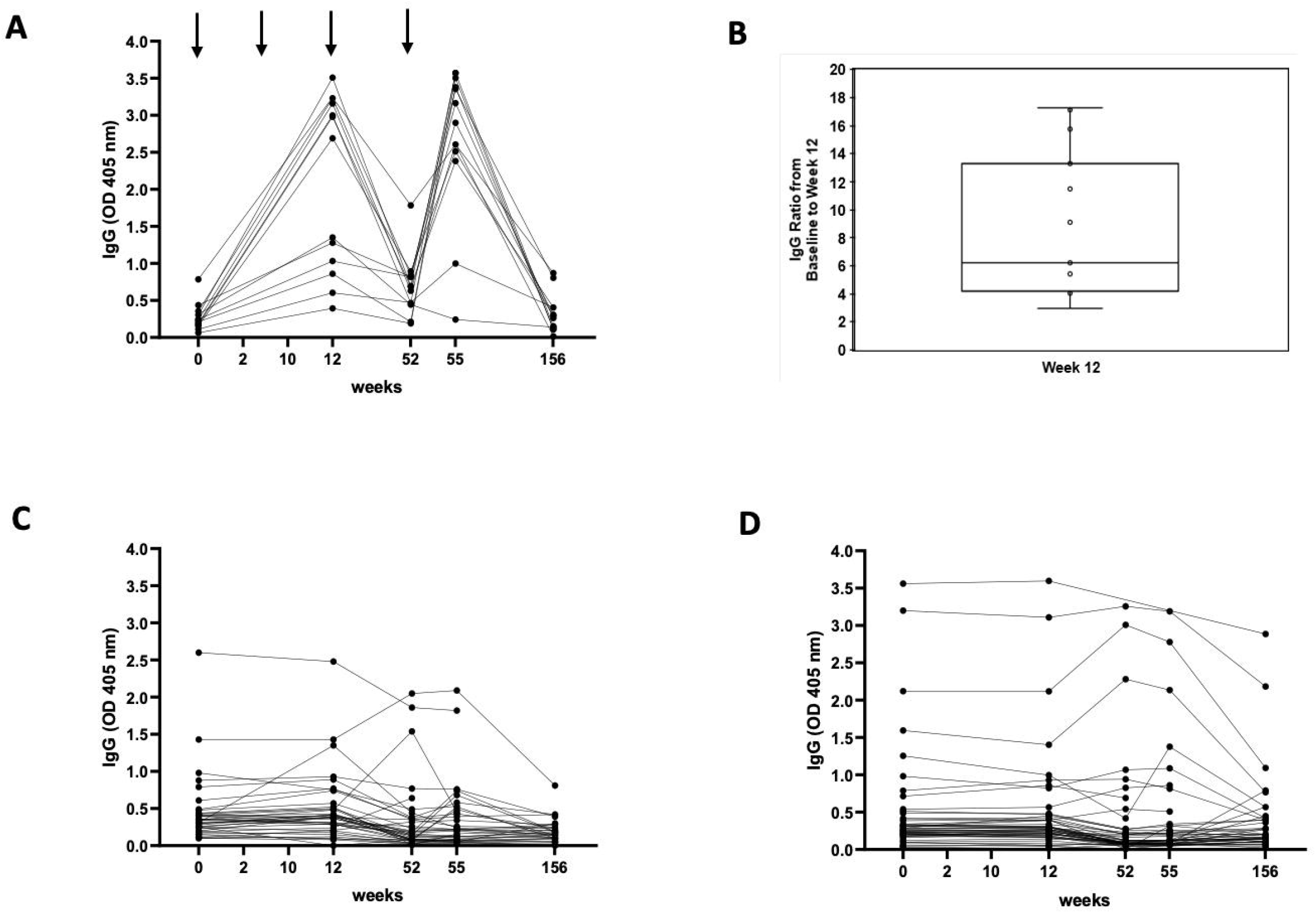
anti-MUC1 IgG Levels: (A) among MUC1 vaccine recipients with an immune response (N=13); (B) Fold elevation in anti MUC1 IgG among vaccine recipients with an immune response; (C) among vaccine recipients without an immune response (N=39); (C) among placebo controls (N=50). Arrows indicate timing of vaccine administration. Blood sampling occurred at the weeks enumerated on the X-axis. Results are shown for 1:80 plasma dilution.

### Correlates of Vaccine Responsiveness

#### Myeloid derived suppressor cells (MDSC)

Three different MDSC subpopulations, M-MDSC, PMN-MDSC and e-MDSC in pre-vaccination PBMC were evaluated in 47 subjects in relation to immune response at week 12 (Table S2). Non-responders (N=34) had a higher mean (±SD) level of circulating PMN-MDSC pre-vaccination than responders (N=13), (0.8 ± 1.1 vs. 0.2 ± 0.09, P=0.0006), and of e-MDSC (0.4 ± 0.3 vs. 0.2 ± 0.11, P=0.02) (Table Supplement S1). There was no association between pre-vaccination levels of M-MDSC and response to the vaccine.

#### Cytokines

Cytokine concentrations in pre-vaccination plasma from those with an immune response at week 12 (N=13) were compared with non-responders (N=39) (Figure 3). Circulating levels of IL-6 (p=0.0194) and IL-8 (p=0.001) were significantly higher in non-responders. Other differences did not reach statistical significance.

**Figure 3:**
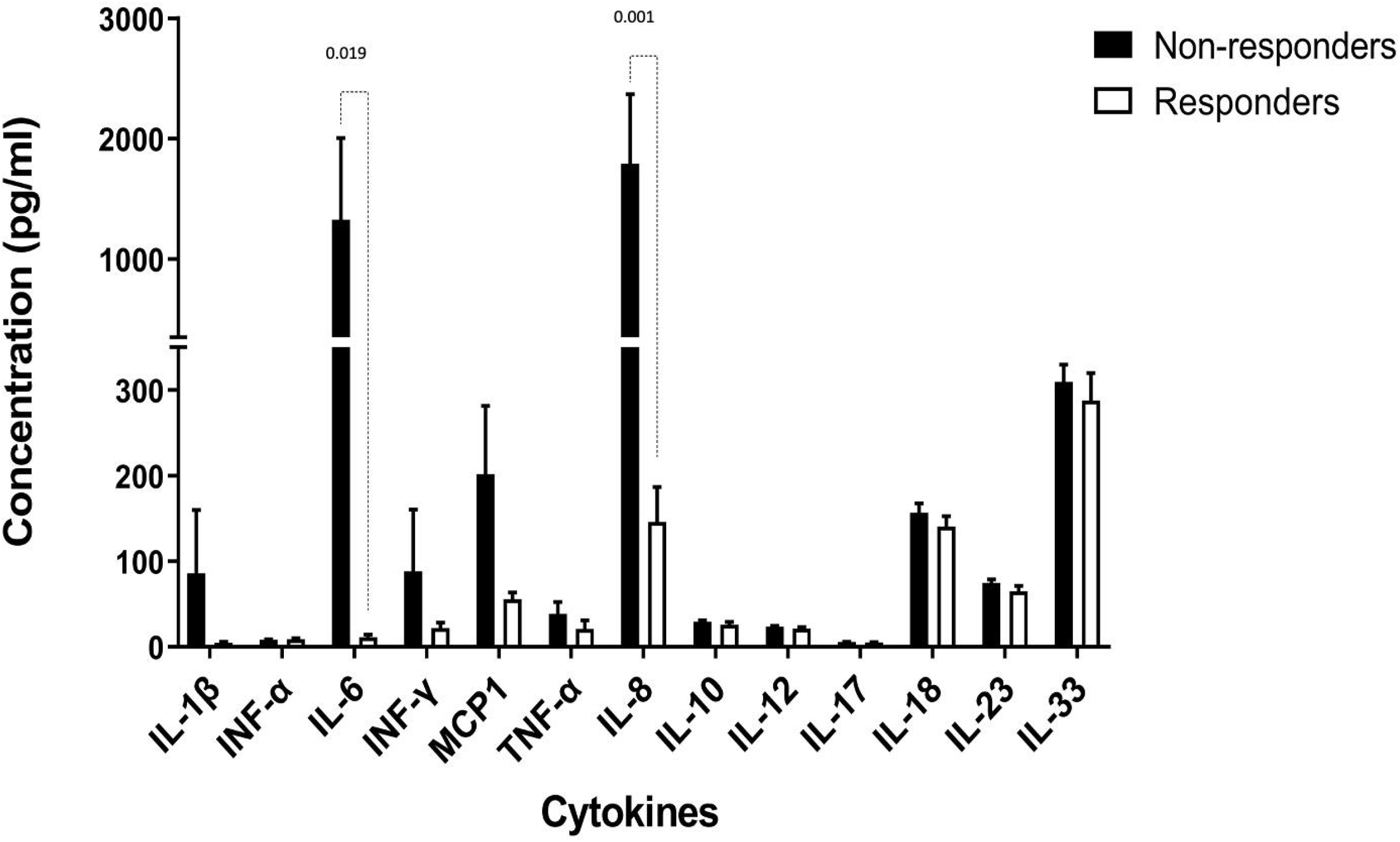
Pre-vaccination levels of circulating proinflammatory cytokines in MUC vaccine. recipients. White bars: vaccine recipients with an immune response (N=13); black bars, vaccine recipients without an immune response (N=39). The Y-axis shows cytokine concentrations measured by bead-based multiplex cytokine assay (LEGENDplex ™, Biolegend, San Diego, CA, USA) at 1:2 plasma dilution.

#### Factors Associated with Immune Response at Week 12

Women were more likely to manifest an immune response to the vaccine (9/20, 45%) than men (4/32, 12.5%, P=.009). In a univariate logistic regression model examining demographic factors, NSAID or aspirin use, adenoma characteristics and MDSC populations, females had higher odds of immune response than males (OR=6.5 [95% CI, 1.6-25.9], P=0.005), and PMN-MDSC and e-MDSC as continuous variables were associated with a lower odds of immune response (OR=0.54 [95% CI, 0.33 - 0.88], P=<0.001), and (OR=0.003 [95% CI, <.001 - 0.486], P=0.002), respectively. In a multivariate logistic regression, only female gender was significantly associated with a higher likelihood of having an immune response (OR=5.79 [95% CI, (1.09 – 30.8)], P=0.04), and a 1 unit increase in PMN-MDSC led to a borderline significant lower odds of immune response (OR=0.004 [95% CI, (<0.001 – 1.55], P=0.07).

#### Adenoma Recurrence

The mean ± SD time to follow up colonoscopy from initial vaccination was 886.1 ± 248.9 days for the MUC1 vaccinated group versus 923.0 ± 258.7 days in the placebo group (P=0.36). The unadjusted and adjusted multivariate relative risk association of various types of adenoma recurrence including any adenoma recurrence, advanced or multiple adenoma recurrence, adenoma recurrence restricted to an adenoma >5mm, and an advanced adenoma recurrence restricted to a different anatomic segment than the baseline advanced adenoma in relation to receipt of vaccine and to being an immune responder are shown in Table 2. A recurrent adenoma was observed in 31 of 47 (66.0%) participants receiving placebo vs. 27 of 48 (56.3%) participants in the MUC1 group (adjusted relative risk (aRR) = 0.83 [95% CI, 0.60-1.14], P=0.25). In immune responders (N=11), any adenoma recurrence was observed in 3 of 11 (27.3%) (aRR= 0.41 [95% CI, 0.15-1.11], P=0.08). When restricting recurrence to adenomas >5mm, the adjusted RR point estimate for recurrence was lower in both the MUC1 group (RR=0.79 [95% CI, 0.46-1.36], P=0.41) and immune responder group (RR=0.17 [95% CI, 0.02-1.11], P=0.06) (Table 2). There was no statistically significant difference in the mean number of adenomas per participant between the MUC1 and the placebo groups (1.0 ± 1.26 vs. 1.5 ± 2.19, respectively; P=0.29), but there was a difference in total number of recurrent adenomas in the placebo group (N=71) vs. the vaccine group (N=47), P=0.05 (Table supplement S2).

**Table 2.** Relative Risk of Recurrent Adenoma(s)

| Table 2. Relative Risk of Recurrent Adenoma(s) |  |  |  |  |  |
| --- | --- | --- | --- | --- | --- |
| Recurrent Adenoma(s) | N with Recurrent<br>Adenoma / Total N (%) | Unadjusted Relative<br>Risk (95% CI) | P-value<sup>1</sup> | Adjusted<sup>2</sup> Relative<br>Risk (95% CI) | P-value<sup>3</sup> |
| **Any adenoma** |  |  |  |  |  |
| Placebo (reference) | 31/47 (66.0%) | 1.00 | - | 1.00 | - |
| MUC1 | 27/48 (56.3%) | 0.85 (0.62-1.18) | 0.3395 | 0.83 (0.60-1.14) | 0.2486 |
| Immune Responder<sup>3</sup> | 3/11 (27.3%) | 0.41 (0.15-1.11) | 0.0791 | 0.41 (0.15-1.11) | 0.0791 |
| **Advanced adenoma** |  |  |  |  |  |
| Placebo (reference) | 4/47 (8.5%) | 1.00 | - | 1.00 | - |
| MUC1 | 7/48 (14.6%) | 1.71 (0.54-5.47) | 0.3693 | 1.69 (0.53-5.39) | 0.3820 |
| Immune Responder<sup>3</sup> | 1/11 (9.1%) | 1.07 (0.13-8.64) | 0.9551 | 0.86 (0.11-7.00) | 0.8995 |
| **Multiple adenomas** |  |  |  |  |  |
| Placebo (reference) | 13/47 (27.7%) | 1.00 | - | 1.00 | - |
| MUC1 | 12/48 (25.0%) | 0.90 (0.46-1.77) | 0.7812 | 0.87 (0.44-1.71) | 0.7004 |
| Immune Responder<sup>3</sup> | 0/11 (0.0%) | NE | - | NE | - |
| **Advanced adenoma, in different**<br>**location from baseline<sup>5</sup>** |  |  |  |  |  |
| Placebo (reference) | 3/47 (6.4%) | 1.00 | - | 1.00 | - |
| MUC1 | 1/48 (2.1%) | 0.33 (0.04-3.03) | 0.3295 | 0.13 (0.01-1.22) | 0.0741 |
| Immune Responder<sup>3</sup> | 0/11 (0.0%) | NE | - | NE | - |
| **Adenoma > 5 mm in size<sup>6</sup>** |  |  |  |  |  |
| Placebo (reference) | 19/47 (40.4%) | 1.00 | - | 1.00 | - |
| MUC1 | 15/48 (31.3%) | 0.77 (0.45-1.33) | 0.3600 | 0.79 (0.46-1.36) | 0.4061 |
| Immune Responder<sup>3</sup> | 1/11 (9.1%) | 0.22 (0.03-1.50) | 0.1239 | 0.17 (0.02-1.11) | 0.0630 |

**Abbreviations:** CI=Confidence Interval; NE = Not Estimable
<sup>1</sup>Wald test
<sup>2</sup>Risk ratios has been adjusted for body mass index, gender, nonsteroidal anti-inflammatory drug use, aspirin use, multiple baseline adenomas, and multiple baseline advanced adenomas
<sup>3</sup>Participants who responded at both Week 12 and Week 55
<sup>4</sup>Participants who did not respond at both Week 12 and Week 55
<sup>5</sup>Excluding advanced adenomas as recurrences for those detected in the same segment of the bowel as baseline advanced Adenomas
<sup>6</sup>Excluding adenomas $\leq 5$ mm as recurrences

The mean PMN-MDSC level within the vaccine group at baseline was higher among those with an adenoma recurrence compared to those without an adenoma recurrence (0.7 ± 0.99) vs. (0.6 ± 1.07), P=0.06. Within the placebo arm, there was no difference in mean PMN-MDSC between those with adenoma recurrence (0.8 ± 0.54) vs. those without adenoma recurrence (1.0 ± 1.59), P=0.65.

We measured levels of anti-MUC1 IgG at the time of follow-up colonoscopy in plasma samples from 76 individuals within ± 4 weeks of the colonoscopy. In the MUC1 group (N=36) the time of blood draw was 922 days ± 196 versus 946 days ± 222 in the placebo group (N=40), P=0.36. Antibody levels were low, not different from baseline, and there was no association with adenoma recurrence. The mean IgG OD ±SD at the time of follow-up colonoscopy was 0.34 ± 0.42 for those that had an adenoma recurrence and 0.31 ± 0.51 for those that did not (P=0.63).

#### Vaccine safety

Grade 1 or higher adverse events (AEs) were more common in vaccine recipients compared with placebo (96.2% vs. 76.0%, respectively; P=0.003), as were grade 2 or higher AEs (84.9% vs. 52.0%, respectively; P=0.0003), primarily due to greater injection site reactions in the MUC1 group. There was no difference in grade 3 or higher AE’s between the MUC1 and the placebo groups (20.8% vs. 20.0%, respectively; P=0.92) and no serious adverse events were attributable to the vaccine. A detailed accounting of all adverse events is provided in supplemental tables S3-S5.

#### Expression of MUC1 and the immune microenvironment of baseline and recurrent adenomas

MUC1 expression by immunohistochemistry, was evaluated in paired baseline and recurrent adenomas in 8 participants, six selected randomly from the placebo group and from two of the three MUC1 vaccine immune responders who recurred. In both groups, there was preserved MUC1 expression in both the baseline and the recurrent adenomas, with extent and intensity of staining rated at the highest value of 4. Localization was predominantly apical and cytoplasmic with no pattern of change (Figure 4).

**Figure 4:**
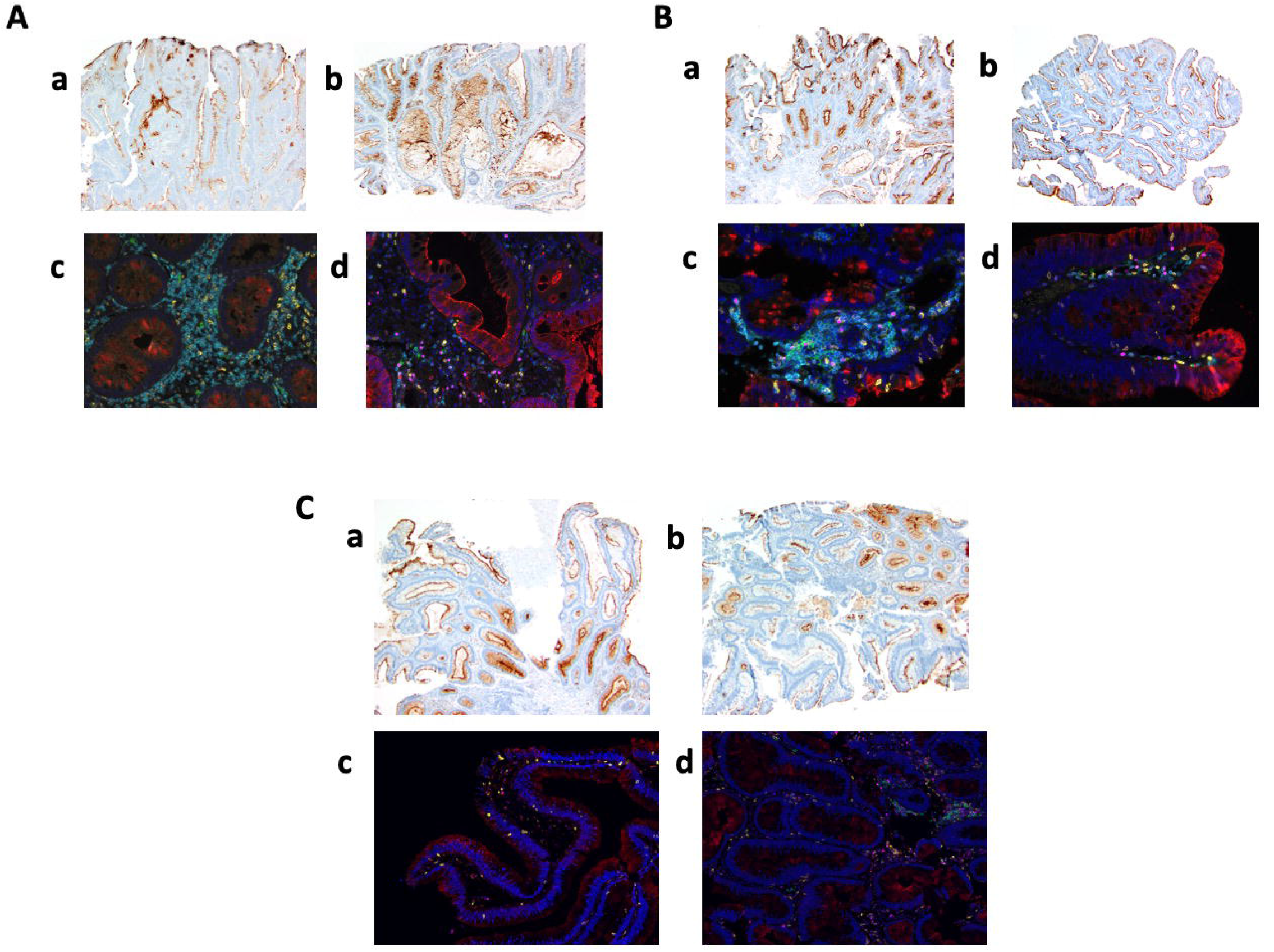
MUC1 expression and immune infiltrate in baseline and recurrent adenoma pairs. Examples are shown of 2 placebo controls (A and B) and one MUC1 responder (C). Anti-MUC1 antibody was used to stain paraffin-embedded tissue sections (panels a and b). Antibodies against CD4 (blue), CD8 (yellow), CD220 (green), FoxP3 (magenta), CD15 (red) and multi-spectral imaging were employed to detect CD4 T cells, CD8 T cells, B cells, regulatory T cells and myeloid cells, respectively (panels c and d).

#### Multi-spectral Imaging

Using multi-spectral imaging, we examined the immune microenvironment of adenomas. We characterized the infiltrating immune cells in 43 matched adenoma pairs (baseline and recurrent), 26 from the placebo group and 17 from the MUC1 vaccinated group (15 non-responders and 2 vaccine responders, though recurrent adenoma was not available from one of the vaccine responders). Figure 5 is a bioinformatic summary of the landscape of immune infiltrates. The cell density score expressed as a heat map demonstrates that the stroma was more heavily infiltrated than the epithelial component in the baseline and the recurrent adenomas. The epithelium of recurrent adenomas was more heavily infiltrated than in the baseline adenomas, primarily due to CD4 T cells that increased in recurrent adenomas in both the placebo (P=0.058) and MUC1 group (P=0.059) (Figure 5B). B cell (CD20) infiltration in the epithelium was low in both groups (Figure 5B). FoxP3 regulatory T cells and CD15 myeloid cell immunosuppressive populations which include MDSCs, were significantly higher in the recurrent adenomas in the placebo group (FoxP3 p=0.044; CD15 p=0.0024), but were not significantly different in the MUC1 non-responder group (Foxp3 p=0.20; CD15 p=0.37).

**Figure 5:**
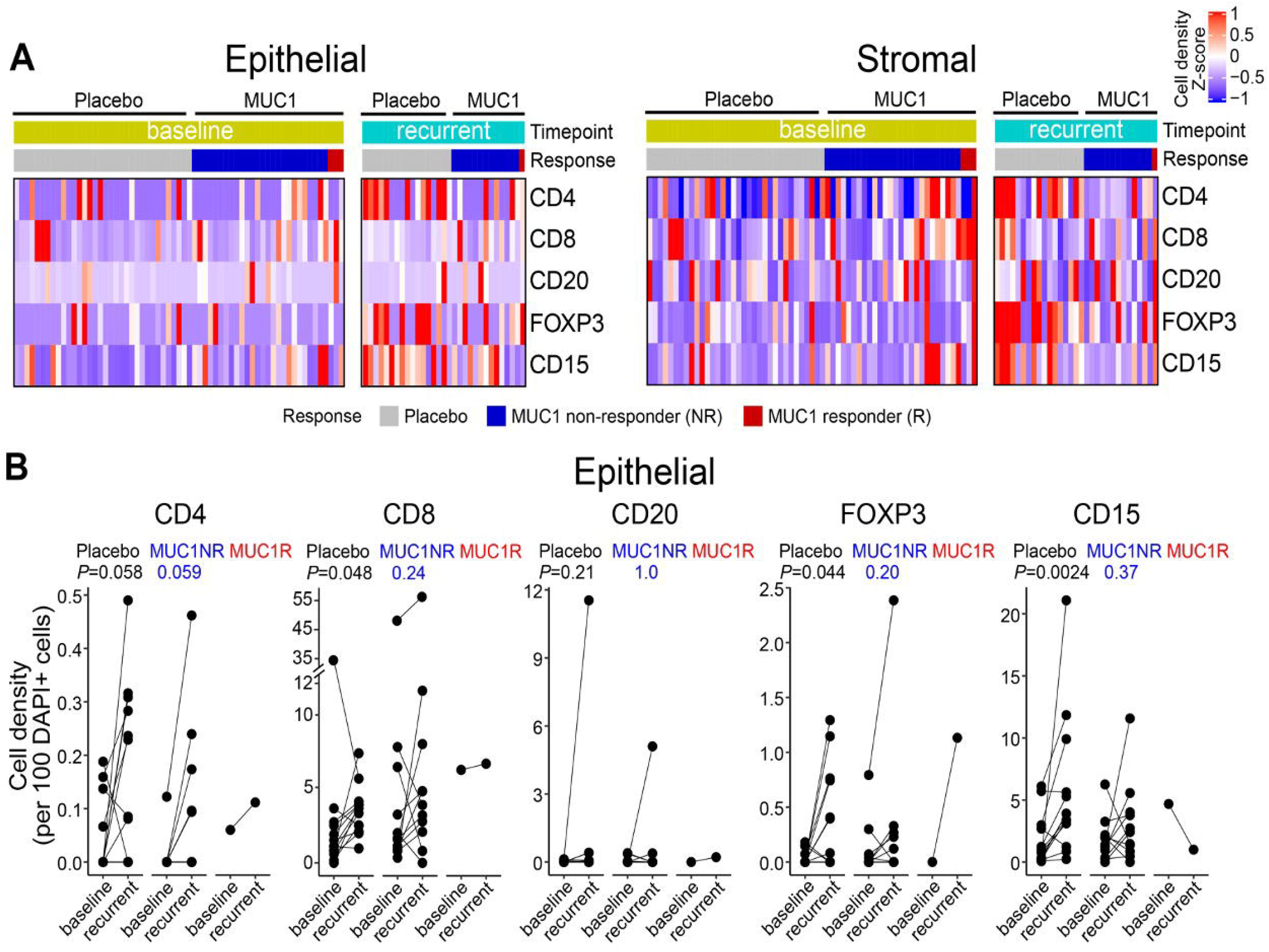
Immune infiltrates in colon adenomas. **(A)** Epithelial and Stromal distribution of CD4^+^ T cells, CD8^+^ T cells, CD20^+^ B cells, FOXP3^+^ regulatory T cells, and CD15^+^ myeloid cells in adenomas. Results are grouped by randomization (MUC1 vaccine or placebo), by baseline or recurrent adenomas, and by immune response to the vaccine. In the heat maps, each row depicts a cell population, and each column represents one trial patient. Data are shown for individuals from the baseline adenoma in placebo group (N=25), baseline adenoma in MUC1 recipients (N=17), recurrent adenoma in placebo group (N=14), and recurrent adenoma in MUC1 group (N=12). Cell density was calculated as the number of marker-positive cells divided by each compartment’s total number of cells. Cell density of each sample across all regions of interest (ROIs) per compartment was collapsed at sample level by median and then collapsed at patient level by median when multiple samples per individual were tested. The dendrogram shows clustering of the samples based on cell density with Euclidean distance. **(B)** Comparison of immune infiltrates in the epithelial compartment between baseline and recurrent adenomas within individuals from the placebo and MUC1 vaccine groups. Data are shown for placebo (N=13), MUC1 non-responder (MUC1NR) (N=11), and MUC1 responder (MUC1R) (N=1). Samples from the baseline and recurrent adenoma within an individual are connected by a line and are compared via a two-sided paired t-test.

## Discussion

This is the first randomized, double-blind, multicenter trial of a vaccine for the prevention of human non-viral neoplasia, based on a shared, non-mutated tumor associated antigen. Twenty-five percent of vaccine recipients developed an immune response at week 12 compared with 0% in the placebo group, a highly significant difference. Using a doubling of anti-MUC1 IgG as a marker of response to both the initial vaccination and a memory response to the booster dose at 1 year, 22% of MUC1 vaccine recipients (11/51) were classified as immune responders. For the clinical outcome of adenoma recurrence, 66% in the placebo group recurred compared with 56% in the vaccine arm, a non-significant difference. Among participants who were immune responders, 27% (3/11) had a recurrent adenoma, a 38% absolute reduction compared to the placebo group (66% recurrence), which approached, but did not reach statistical significance (P=0.08).

In a pilot study of this same vaccine in individuals with advanced adenoma, administered in a similar fashion and schedule (11), there was a 44% (17/39) response at week 12 compared to 25% in this trial. The smaller than expected percentage of participants who met criteria for being an immune responder in this randomized trial limited the power of the study to demonstrate a significant effect on the clinical efficacy endpoint of adenoma recurrence.

In terms of the magnitude of the immune response, in the pilot study we observed an increase in the ratio of anti-MUC1 IgG at week 12 of up to 36-fold with a mean ratio increase among responders of 13.5, whereas in this study the ratio increase went to 17-fold with a mean ratio increase among responders of 8.9 (Figure 2B).

The reasons why the response rate to the initial vaccination and the magnitude of the immune response were reduced in this trial compared to the pilot study are unclear. One potentially important difference between the two studies was the time between the advanced adenoma removal and the administration of the vaccine. In the pilot study, participants could have had an advanced adenoma at any time prior to enrollment, and some had their adenoma removed up to 9 years prior. In the current study, vaccine administration occurred within 1 year of adenoma removal.

The multispectral images of the immune infiltrate in the baseline advanced adenomas demonstrate a presence of suppressive regulatory T cells (FoxP3^+^ Tregs) and MDSC (CD15^+^). Evidence is beginning to accumulate that these cells can suppress systemic as well as local immune responses (29-31). Even though the baseline adenoma was removed prior to vaccination, Tregs could remain in the circulation and MDSCs could continue to be generated due to the continuous presence of proinflammatory cytokines produced by both innate and adaptive immune recognition.

Evidence of continued systemic immunosuppression post-adenoma removal is demonstrated in our analysis of serum cytokines. IL-6 and IL-8 in particular are proinflammatory cytokines that are biomarkers of ongoing inflammation and known to negatively affect adaptive immune responses. They have been shown previously to be elevated in the sera of cancer patients (32, 33) and to modulate response to immunotherapy (34, 35). More recently they have also been found in premalignant disease, such as in individuals with cervical epithelial neoplasms (36), oral premalignant lesions (37, 38), and Barrett’s esophagus (39). The increased levels of these cytokines in non-responders compared to responders suggests that even in the setting of premalignancy, vaccines may need to be combined with other non-toxic approaches to modulating immunosuppression.

In some individuals the baseline levels of anti-MUC1 antibodies were relatively high and did not increase after vaccination. One reason why those with high antibody titers at baseline did not respond to the vaccine could be that the pre-existing antibodies cleared the administered vaccine antigen, preventing its presentation and facilitation of a memory response. Alternatively, the antibodies at baseline were generated when the individuals were immunocompetent but immunosuppressive mechanisms prevented a renewed response to the vaccine. We did not observe a correlation between high baseline anti-MUC1 antibody levels and adenoma recurrence, but we did observe a lower adenoma recurrence rate in those who generated both an initial and a memory response.

We reproduced the observation made in the pilot study that circulating MDSCs, but not circulating Tregs, were an immune correlate of vaccine non-responsiveness. The availability of the placebo group in this trial, allowed us to distinguish between the MDSC role in suppressing immune responses to the vaccine, versus their role in preventing adenoma recurrence. MDSC levels were similar in placebo participants with and without adenoma recurrence. We do not know why some individuals with advanced adenoma have higher levels of circulating MDSC. Response to the vaccine is compromised by the presence of these cells. PMN-MDSC levels could be used to select participants who would be more likely to respond to the MUC1 vaccine. Development of safe and effective approaches for MDSC neutralization of their immunosuppressive effect are also potential considerations. Supplemental arginine, an amino acid important for T cell survival (40), could replace arginine depleted by the MDSC-produced arginase (41). Phosphodiesterase inhibitors inhibit MDSC (42), as does priming with systemic poly IC-LC (19). Administration of cytokines such as IL-7 and IL-15 could promote T cell functioning (43).

Women were more likely to respond to the vaccine than men. Sex differences in response to vaccination and various immunotherapies have been previously observed and explanations proposed (44-47). MUC1 vaccine may be more immunogenic in females than in males because females experience a larger number of epithelial inflammatory events such as ovulatory cycles, endometriosis, and use of intrauterine devices, that affect epithelial tissues and transiently change MUC1 expression from normal to the tumor-like form, which can lead to spontaneous, albeit low level, immune priming and immune memory for abnormal MUC1 (48, 49). This immune memory could facilitate the response to the MUC1 vaccine resulting in a stronger immune response.

We used anti-MUC1 IgG as the marker for response to the vaccine. IgG is a highly stable molecule that is simple and cost effective to measure. Because MUC1-specific B cells require MUC1-specific T cell help to produce IgG, this is also an indirect measure of the T cell response. MUC1 is a cell surface antigen and anti-MUC1 antibodies could be important effector molecule for elimination of epithelial tissue expressing tumor-associated MUC1 through antibody dependent cellular cytotoxicity (ADCC) and antibody-dependent cellular phagocytosis (ADCP) (50). Tumor (or a premalignant lesion) destruction by ADCC and ADCP results in antigen release and antigen presenting cell activation, leading to increased antigen presentation and generation of new immune responses, known as epitope spreading (51). Inducing immunity to one immunogenic antigen can create a favorable environment for the generation of immune responses to other antigens that may have an even stronger protective effect.

Considering that use of non-mutated tumor-associated antigens as immunotherapy can potentially generate autoimmunity (52), it is notable that the MUC1 vaccine was well tolerated without significant toxicity other than injection site reactions that resolved without treatment. To confirm the tumor MUC1 specificity of the vaccine-elicited MUC1 immunity, we cloned multiple anti-MUC1 IgG antibodies from a vaccine responder and demonstrated that they reacted with MUC1 on tumor cells and not on healthy tissues (53). Thus, autoimmunity and other toxicities were not a limiting factor. Most importantly, we demonstrated the feasibility of testing tumor-associated antigen vaccines in the preventive setting, permitting an understanding of vaccine immunogenicity and safety without the confounding issues of cancer and cancer therapy. Our study demonstrates that individuals at increased risk for cancer are amenable to participating in vaccine trials to reduce their risk for cancer.

In this placebo controlled, randomized trial of MUC1 vaccine, immune responses to MUC1 were limited to vaccine recipients. There was a non-significant, but suggestive reduction in adenoma recurrence amongst those with a significant immune response. Efforts to further improve the immune response to the vaccine are needed to better assess its potential for colorectal cancer prevention.

## Data Availability

All data produced in the present study are available upon reasonable request to the authors

