## supplemental appendix for "Randomized, Double-Blind, Placebo-Controlled Trial of MUC1 Peptide Vaccine for Prevention of Recurrent Colorectal Adenoma"

**Contents**

Methods Supplement M1: Trial Administration	Page 2

Methods Supplement M2: Detailed Laboratory Methods	Page 3-5

Figure S1: Endpoint titers of plasma antibodies amongst vaccine responders	Page 6

Table S1: MDSC values by vaccine immune response vs. non-response	Page 7

Table S2: Number of Adenomas at Endpoint Colonoscopy	Page 8

Table S3: Grade 1 Adverse Events	Page 9-11

Table S4: Grade 2 Adverse Events	Page 12-14

Table S5: Grade 3 Adverse Events	Page 15

**Methods Supplement M1: Trial Administration**

Vaccine:

The MUC1 vaccine consisted of a 100-amino acid synthetic MUC1 peptide with the amino acid sequence corresponding to 5 tandem repeats of 20 amino acids in length, derived from the extracellular domain known as the Variable Number of Tandem Repeats (VNTR) region. The peptide H_2_N-(GVTSAPDTRPAPGSTAPPAH)_5_-CONH_2_ was synthesized at the University of Pittsburgh Clinical Peptide Synthesis Facility.

The vaccine consisted of 100µg of the MUC1 100mer synthetic peptide dissolved in 50µL of sterile saline, admixed with 500µg of 250µL Hiltonol® for a total injection volume of 300µL. Placebo injection was 300µL of sterile saline. Vaccine or placebo was administered subcutaneously, blinded to content, in the upper thigh, and when possible, in the same upper thigh at each injection. The trial received IRB approval at each site under an Investigational New Drug (IND) approval from the FDA. It was registered at ClinicalTrial.gov under NCT-007773097.

Adverse Events:

Laboratory monitoring included CBC and liver function tests performed at baseline, prior to each vaccine dose, and at week 55. BUN and creatinine were evaluated at baseline and at week 52. A repeat ANA test was performed at week 52 prior to the booster vaccination. Injection site reactions were monitored using a participant-completed vaccine report card, with classification of none, mild, moderate and severe reaction based on impairment to physical activity, and dimension of erythema and swelling.

**Methods Supplement M2:** **Detailed Laboratory Methods**

**Measurement of Anti-MUC1 IgG, Cytokines and Myeloid-Derived Suppressor Cells (MDSC)**

Heparinized blood, shipped to the University of Pittsburgh, was layered on lymphocyte separation medium (MPbio) and centrifuged at 800 g for 10 min with lowest acceleration and deceleration speed, within 24 hours from when it was drawn. Plasma was collected from the top of the separation tube and frozen in small aliquots at −20°C. PBMC were collected from the interphase between plasma and separation medium, washed twice, re-suspended in 80% human serum and 20% DMSO, and stored in liquid nitrogen at -80^o^C.

**Anti MUC1 IgG**

Enzyme-linked Immunosorbent Assay (ELISA) was used to measure plasma anti-MUC1 IgG levels. 1 µg/ml of the 100mer MUC1 vaccine peptide dissolved in 2.5% bovine serum albumin (BSA) in PBS was coated onto Immulon 4HBX ELISA plates (Thermo-Fisher Scientific, MA) in triplicate wells. Triplicate wells coated with 2.5% bovine serum albumin (BSA) in PBS served as no antigen controls for non-specific binding. Plates were placed overnight at 4 °C. Plasma was serially diluted from 1:40 to 1:640 in 2.5% bovine serum albumin (BSA), added to ELISA plates, and kept for 1 hour at room temperature. Plates were washed five times with 0.1% Tween 20 detergent in PBS. 50µl alkaline phosphatase conjugated with anti-human IgG (Sigma-Aldrich) in 2.5% BSA DPBS was added, and the plate incubated for 1 hour at RT. The plate was washed again, 100 ul of p-nitrophenyl phosphate (Sigma-Aldrich) added and the plate incubated for 1 hour in the dark. The reaction was stopped with 50µl 0.5M NaOH. The plates were read at OD405nm on the spectrophotometer SpectraMaxi3 (Molecular Devices, San Jose, CA, USA). Control (no antigen) plate was put through the same reactions except that 50µl PBS were added instead of the MUC1 peptide. OD values from the no antigen wells were subtracted from corresponding values on the antigen-coated wells.

**Myeloid-Derived Suppressor Cells (MDSC)**

MDSC subpopulations were characterized based on their cell surface markers into polymorphonuclear (PMN)-MDSC, CD11b^+^HLA-DR^−/low^ CD33^+^ CD15^+^ CD14^-^, monocytic (M)-MDSC: CD11b^+^HLA-DR^−/low^ CD33^+^ CD15^−^ CD14^+^ and early (e)-MDSC:CD11b^+^HLA-DR^-/low^ CD33^+^ CD15^-^ CD14^-^. Frozen PBMC were thawed in the 37°C water bath, washed, and stained with APC labeled anti-human CD11b (BD Biosciences Clone:ICRF44), PE-Texas/Red labeled anti-human CD33 (BD Biosciences Clone:WM53), FITC labeled anti-human HLA-DR (BD Biosciences Clone:G46-6), V450 labeled Anti-human CD14 (BD Biosciences Clone:MϕP9) and PE-Cy7 labeled anti human CD15 (BD Biosciences Clone:HI98). Stained cells were analyzed on IMM Fortessa (BD Bioscience) and data analyzed using FlowJo (v10) software (FlowJo LLC).

**Cytokines**

A 1:2 dilution of plasma was used for the determination of cytokine concentrations. Bead-based multiplex cytokine assay was used to measure IL-1β, INF-α, IL-6, INF-γ, MCP1, TNF-α, IL-8, IL-10, IL-12, IL-17, IL-18, IL-23, IL-33 (LEGENDplex^TM^, Biolegend, San Diego, CA, USA) (Supplement S1). The cytokines in this panel were chosen based on their previously reported roles in inflammation and cancer (32, 33). IL-1β, INF-α, IL-6, INF-γ, MCP1, TNFα and IL-8 (or CXCL8) are proinflammatory cytokines that are known to suppress immunity and IL-10 can suppress both innate cell-mediated and adaptive immunity. IL-12, IL-17, IL-18 and IL-23 promote type 1 immunity while IL-33 promotes type 2 immunity (34). Serial dilution of the cytokine panel was run on the same plate according to manufacturer’s instructions ((LEGENDplex^TM^, Biolegend, San Diego, CA, USA) and read using a Fortessa flow cytometer. Ten thousand total events were recorded per plasma sample and cytokines were considered absent or undetectable if below 2 pg/ml concentration.

**Immunohistochemistry (IHC) Staining of Polyps for MUC1 Expression**

Four-micron thick formalin-fixed, paraffin embedded adenoma sections were deparaffinized in xylene, and rehydrated through graded alcohols to distilled water before undergoing antigen retrieval using the Ventana (Tucson, AZ) CC1 protocol (36 minutes at 95^o^ C).  The sections were stained using a rabbit monoclonal antibody against MUC1 (1:100 dilution, clone EPR1023, catalog number ab109185, Abcam, Cambridge MA) as a primary antibody, with a 16-minute incubation time at 36^o^ C using the Ventana BenchMark Ultra (Roche Diagnostics). Stained tissues were evaluated for the extent of staining (0=no staining, 1=1-10%, 2=11-25%, 3=26-40%, 4=>40%), intensity of staining (0-4+), and localization of MUC1 (apical versus cytoplasmic).

**Multiplex immunofluorescence staining**

Uniplex immunofluorescence staining was performed manually using the Opal 6-Plex kit (Akoya Biosciences, Marlborough, MA), which uses individual tyramide signal amplification (TSA)-conjugated fluorophores to detect targets. The slides were scanned using the Vectra Polaris spectral imaging system (Akoya Biosciences) according to previously published instructions (36).

After deparaffinization, slides were placed in a plastic container filled with antigen retrieval (AR) buffer in Tris-EDTA buffer and subjected to boiling (1 min) at 100 °C. The sections were then microwaved for an additional 15 min at 75 °C. Slides were allowed to cool in the AR buffer for 15 min at room temperature and were then rinsed with deionized water and 1 × Tris-buffered saline with Tween 20. To initiate protein stabilization and background reduction, Tris-HCl buffer containing 0.1% Tween was used for 10 min at room temperature. Slides were then incubated between 30 min and 2 h with antibodies against the immune markers at specific dilutions: a-CD20 (dilution 1:50, Abcam, Waltham, MA), a-CD8 (dilution 1:10, Abcam), a-CD15 (dilution 1:25, Abcam), a-FOXP3 (dilution 1:10, Spring Biosciences, Pleasanton, CA), and CD4 (dilution 1:1, Biocare Medical, Pacheco, CA). Finally, the slides were washed and incubated for 10 min at room temperature with secondary antibodies (Novocastra, Leica Biosystems) after successive washes in TBST.

The slides were then incubated at room temperature for 10 min with one of the following Alexa Fluor tyramides (Akoya Biosciences) included in the Opal 6 Kit to detect antibody staining, prepared according to the manufacturer’s instructions: Opal 520 (CD20), Opal 540 (CD8), Opal 570 (CD15), Opal 620 (FOXP3), and Opal 650 (CD4) (dilution 1:50). After three additional washes in deionized water, the slides were counterstained with DAPI for 5 min and mounted with VECTASHIELD Hard Set (Vector Labs, Burlingame, CA). Autofluorescence (negative control) slides were also included, using primary and secondary antibodies and omitting the fluor tyramides.

A total of 466 regions of interest (ROI) were identified using Phenochart under the supervision of a gastrointestinal-trained pathologist (ADS). Spectral libraries using the InForm 2.4 image analysis software were then analyzed and trained to separate out the polyp and stroma compartments through tissue segmentation. Five marker-positive cells were annotated, including CD4+ T cells, CD8+ T cells, CD15+ myeloid cells, CD20+ B cells, and FOXP3+ regulatory T cells. In total, 1,032,442 segmented cells with classified phenotypes were included.

**Image collection and analysis**

Multiplex immunofluorescence staining was imaged using the fluorescence protocol at 10 nm λ from 420 nm to 720 nm, to extract fluorescent intensity information from the slides. A total of 466 regions of interest (ROI) of 0.669 × 500 µm, 0.3345 mm^2^ each were identified using Phenochart under the supervision of a gastrointestinal-trained pathologist (ADS). Spectral libraries using the InForm 2.4 image analysis software were then analyzed and trained to separate out the polyp and stroma compartments through tissue segmentation. Five marker-positive cells were annotated, including CD4+ T cells, CD8+ T cells, CD15+ myeloid cells, CD20+ B cells, and FOXP3+ regulatory T cells. In total, 1,032,442 segmented cells with classified phe

notypes were included in further analysis.

**Figure S1: Endpoint titers of plasma antibodies at week 12 amongst vaccine responders**

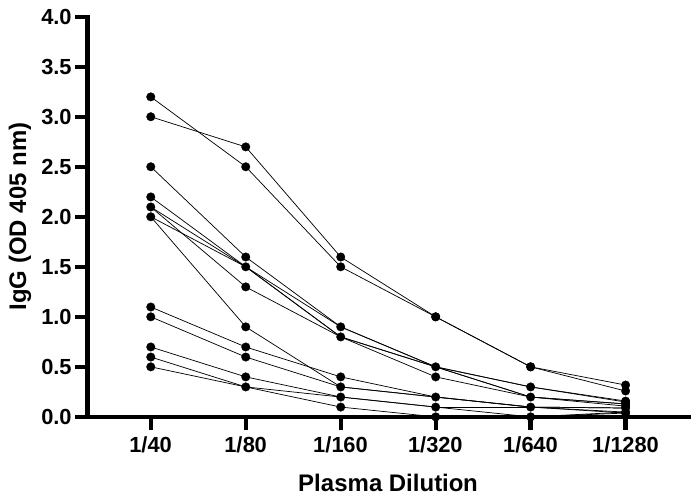

| **MDSC values by Vaccine Immune Response vs. Non-Response** | | | | |
| --- | --- | --- | --- | --- |
|  | Non-  Response (N=34) | Immune Response (N=13) | Total (N=47) | P-value |
| **PMN-MDSC** |  |  |  | **0.0004**^1^ |
| Mean (SD) | 0.8 (1.11) | 0.2 (0.09) | 0.6 (0.99) |  |
| Median | 0.5 | 0.2 | 0.3 |  |
| IQR | 0.3, 0.8 | 0.1, 0.2 | 0.2, 0.7 |  |
| Range | 0.0, 5.0 | 0.0, 0.4 | 0.0, 5.0 |  |
| **M-MDSC** |  |  |  | 0.8864^1^ |
| Mean (SD) | 1.6 (1.14) | 2.2 (2.33) | 1.8 (1.56) |  |
| Median | 1.3 | 1.5 | 1.3 |  |
| IQR | 0.8, 2.4 | 0.6, 3.1 | 0.8, 2.6 |  |
| Range | 0.2, 3.8 | 0.0, 7.4 | 0.0, 7.4 |  |
| **e-MDSC** |  |  |  | **0.0156**^1^ |
| Mean (SD) | 0.4 (0.30) | 0.2 (0.11) | 0.3 (0.28) |  |
| Median | 0.3 | 0.2 | 0.2 |  |
| IQR | 0.1, 0.6 | 0.1, 0.2 | 0.1, 0.4 |  |
| Range | 0.0, 1.1 | 0.0, 0.4 | 0.0, 1.1 |  |
| ^1^Wilcoxon rank sum test | | | | |

**Table S1: MDSC values by vaccine immune response vs. non-response**

|  |  |  |  |
| --- | --- | --- | --- |
| **Number of Subjects (N, %) with Adenomas at Endpoint Colonoscopy** | | | |
|  | **MUC1 (N=48)** | **Placebo (N=47)** | **Total**  **(N=95)** |
| **Number of adenomas at recurrence** ^1^ |  |  |  |
| 0 | 21 (43.8%) | 16 (34.0%) | 37 (38.9%) |
| 1 | 15 (31.3%) | 18 (38.3%) | 33 (34.7%) |
| 2 | 9 (18.8%) | 3 (6.4%) | 12 (12.6%) |
| ≥ 3 | 3 (6.3%) | 10 (21.3%) | 13 (13.7%) |
| Total N of adenomas | 47 | 71 | 118 |
| Mean (SD)^2^ | 1.0 (1.26) | 1.5 (2.19) | 1.2 (1.79) |
| Median | 1.0 | 1.0 | 1.0 |
| IQR | 0.0, 1.5 | 0.0, 2.0 | 0.0, 2.0 |
| Range | 0.0, 6.0 | 0.0, 12.0 | 0.0, 12.0 |
| **Abbreviations:** SD=Standard Deviation; IQR=Interquartile Range  ^1^Fisher’s exact p-value = 0.0546  ^2^Wilcoxon rank-sum p-value = 0.2850 | | | |

**Table S2: Number of Adenomas at Endpoint Colonoscopy**

Table S3: Grade 1 Adverse Events

|  | | **Arm** | |  |  |
| --- | --- | --- | --- | --- | --- |
| **MedDRA (Medical Dictionary for Regulatory Activities) SOC (system organ classes) (v12.0)** | **CTCAE (Common Terminology Criteria for Adverse Events)**  **Term (v4.0)** | MUC1 (N=53) | Placebo (N=50) | Total (N=103) | P-value^1^ |
| Blood and lymphatic sys disorders | Anemia | 1 (1.9%) | 2 (4.0%) | 3 (2.9%) | 0.6104 |
|  | Leukocytosis | 0 (0.0%) | 1 (2.0%) | 1 (1.0%) | 0.4854 |
| Cardiac disorders | Chest pain - cardiac | 2 (3.8%) | 1 (2.0%) | 3 (2.9%) | 1.0000 |
|  | Heart failure | 0 (0.0%) | 1 (2.0%) | 1 (1.0%) | 0.4854 |
|  | Myocardial infarction | 0 (0.0%) | 1 (2.0%) | 1 (1.0%) | 0.4854 |
| Ear and labyrinth disorders | Ear and labyrinth disorders - Oth spec | 0 (0.0%) | 1 (2.0%) | 1 (1.0%) | 0.4854 |
|  | Middle ear inflammation | 1 (1.9%) | 0 (0.0%) | 1 (1.0%) | 1.0000 |
|  | Tinnitus | 0 (0.0%) | 1 (2.0%) | 1 (1.0%) | 0.4854 |
|  | Vestibular disorder | 0 (0.0%) | 1 (2.0%) | 1 (1.0%) | 0.4854 |
| Endocrine disorders | Hypothyroidism | 0 (0.0%) | 1 (2.0%) | 1 (1.0%) | 0.4854 |
| Eye disorders | Cataract | 1 (1.9%) | 2 (4.0%) | 3 (2.9%) | 0.6104 |
|  | Eye disorders - Other, specify | 1 (1.9%) | 0 (0.0%) | 1 (1.0%) | 1.0000 |
|  | Eyelid function disorder | 0 (0.0%) | 1 (2.0%) | 1 (1.0%) | 0.4854 |
| Gastrointestinal disorders | Abdominal pain | 2 (3.8%) | 1 (2.0%) | 3 (2.9%) | 1.0000 |
|  | Bloating | 1 (1.9%) | 1 (2.0%) | 2 (1.9%) | 1.0000 |
|  | Colitis | 1 (1.9%) | 0 (0.0%) | 1 (1.0%) | 1.0000 |
|  | Colonic obstruction | 1 (1.9%) | 0 (0.0%) | 1 (1.0%) | 1.0000 |
|  | Diarrhea | 4 (7.5%) | 1 (2.0%) | 5 (4.9%) | 0.3637 |
|  | Dyspepsia | 3 (5.7%) | 0 (0.0%) | 3 (2.9%) | 0.2433 |
|  | Esophagitis | 1 (1.9%) | 0 (0.0%) | 1 (1.0%) | 1.0000 |
|  | Gastric hemorrhage | 1 (1.9%) | 0 (0.0%) | 1 (1.0%) | 1.0000 |
|  | Gastroesophageal reflux disease | 0 (0.0%) | 2 (4.0%) | 2 (1.9%) | 0.2332 |
|  | Gastrointestinal disorders - Oth spec | 7 (13.2%) | 3 (6.0%) | 10 (9.7%) | 0.3209 |
|  | Gastrointestinal pain | 1 (1.9%) | 0 (0.0%) | 1 (1.0%) | 1.0000 |
|  | Hemorrhoids | 1 (1.9%) | 0 (0.0%) | 1 (1.0%) | 1.0000 |
|  | Nausea | 3 (5.7%) | 1 (2.0%) | 4 (3.9%) | 0.6182 |
|  | Pancreatitis | 1 (1.9%) | 0 (0.0%) | 1 (1.0%) | 1.0000 |
|  | Rectal hemorrhage | 1 (1.9%) | 1 (2.0%) | 2 (1.9%) | 1.0000 |
|  | Rectal pain | 1 (1.9%) | 0 (0.0%) | 1 (1.0%) | 1.0000 |
|  | Small intestinal obstruction | 1 (1.9%) | 0 (0.0%) | 1 (1.0%) | 1.0000 |
|  | Stomach pain | 1 (1.9%) | 0 (0.0%) | 1 (1.0%) | 1.0000 |
|  | Vomiting | 2 (3.8%) | 0 (0.0%) | 2 (1.9%) | 0.4955 |
| General disorders and administration site conditions | Edema limbs | 2 (3.8%) | 0 (0.0%) | 2 (1.9%) | 0.4955 |
|  | Fatigue | 2 (3.8%) | 0 (0.0%) | 2 (1.9%) | 0.4955 |
|  | Fever | 0 (0.0%) | 1 (2.0%) | 1 (1.0%) | 0.4854 |
|  | Flu like symptoms | 5 (9.4%) | 3 (6.0%) | 8 (7.8%) | 0.7164 |
|  | Injection site reaction | 47 (88.7%) | 10 (20.0%) | 57 (55.3%) | **<.0001** |
|  | Non-cardiac chest pain | 0 (0.0%) | 1 (2.0%) | 1 (1.0%) | 0.4854 |
| Hepatobiliary disorders | Hepatobiliary disorders - Other, specify | 1 (1.9%) | 0 (0.0%) | 1 (1.0%) | 1.0000 |
| Immune system disorders | Autoimmune disorder | 0 (0.0%) | 1 (2.0%) | 1 (1.0%) | 0.4854 |
| Infections and infestations | Bronchial infection | 1 (1.9%) | 1 (2.0%) | 2 (1.9%) | 1.0000 |
|  | Conjunctivitis | 0 (0.0%) | 1 (2.0%) | 1 (1.0%) | 0.4854 |
|  | Eye infection | 0 (0.0%) | 1 (2.0%) | 1 (1.0%) | 0.4854 |
|  | Infections and infestations - Oth spec | 3 (5.7%) | 2 (4.0%) | 5 (4.9%) | 1.0000 |
|  | Mucosal infection | 0 (0.0%) | 1 (2.0%) | 1 (1.0%) | 0.4854 |
|  | Nail infection | 2 (3.8%) | 1 (2.0%) | 3 (2.9%) | 1.0000 |
|  | Otitis media | 0 (0.0%) | 1 (2.0%) | 1 (1.0%) | 0.4854 |
|  | Paronychia | 1 (1.9%) | 0 (0.0%) | 1 (1.0%) | 1.0000 |
|  | Sinusitis | 4 (7.5%) | 2 (4.0%) | 6 (5.8%) | 0.6789 |
|  | Skin infection | 1 (1.9%) | 0 (0.0%) | 1 (1.0%) | 1.0000 |
|  | Soft tissue infection | 1 (1.9%) | 0 (0.0%) | 1 (1.0%) | 1.0000 |
|  | Upper respiratory infection | 2 (3.8%) | 0 (0.0%) | 2 (1.9%) | 0.4955 |
|  | Urinary tract infection | 1 (1.9%) | 2 (4.0%) | 3 (2.9%) | 0.6104 |
| Injury, poisoning and procedural complications | Bruising | 0 (0.0%) | 1 (2.0%) | 1 (1.0%) | 0.4854 |
|  | Fall | 2 (3.8%) | 0 (0.0%) | 2 (1.9%) | 0.4955 |
|  | Fracture | 1 (1.9%) | 1 (2.0%) | 2 (1.9%) | 1.0000 |
|  | Inj, pois and proced complic - Oth spec | 2 (3.8%) | 0 (0.0%) | 2 (1.9%) | 0.4955 |
|  | Postoperative hemorrhage | 1 (1.9%) | 0 (0.0%) | 1 (1.0%) | 1.0000 |
|  | Wound complication | 1 (1.9%) | 0 (0.0%) | 1 (1.0%) | 1.0000 |
| Investigations | Alanine aminotransferase increased | 4 (7.5%) | 0 (0.0%) | 4 (3.9%) | 0.1183 |
|  | Alkaline phosphatase increased | 1 (1.9%) | 1 (2.0%) | 2 (1.9%) | 1.0000 |
|  | Aspartate aminotransferase increased | 4 (7.5%) | 0 (0.0%) | 4 (3.9%) | 0.1183 |
|  | Cholesterol high | 2 (3.8%) | 1 (2.0%) | 3 (2.9%) | 1.0000 |
|  | Investigations - Other, specify | 1 (1.9%) | 2 (4.0%) | 3 (2.9%) | 0.6104 |
|  | Lymphocyte count decreased | 1 (1.9%) | 1 (2.0%) | 2 (1.9%) | 1.0000 |
|  | Platelet count decreased | 1 (1.9%) | 0 (0.0%) | 1 (1.0%) | 1.0000 |
|  | Weight loss | 1 (1.9%) | 0 (0.0%) | 1 (1.0%) | 1.0000 |
| Metabolism and nutrition disorders | Hyperglycemia | 2 (3.8%) | 4 (8.0%) | 6 (5.8%) | 0.4280 |
|  | Hyperkalemia | 0 (0.0%) | 1 (2.0%) | 1 (1.0%) | 0.4854 |
|  | Metabolism, nutrition disord - Oth spec | 1 (1.9%) | 0 (0.0%) | 1 (1.0%) | 1.0000 |
| Musculoskeletal and connective tissue disorders | Arthralgia | 5 (9.4%) | 2 (4.0%) | 7 (6.8%) | 0.4380 |
|  | Arthritis | 4 (7.5%) | 3 (6.0%) | 7 (6.8%) | 1.0000 |
|  | Back pain | 9 (17.0%) | 2 (4.0%) | 11 (10.7%) | 0.0527 |
|  | Joint range of motion decr cerv spine | 1 (1.9%) | 0 (0.0%) | 1 (1.0%) | 1.0000 |
|  | Joint range of motion decreased | 1 (1.9%) | 0 (0.0%) | 1 (1.0%) | 1.0000 |
|  | Musculoskeletal, conn tissue - Oth spec | 4 (7.5%) | 2 (4.0%) | 6 (5.8%) | 0.6789 |
|  | Myalgia | 1 (1.9%) | 1 (2.0%) | 2 (1.9%) | 1.0000 |
|  | Neck pain | 1 (1.9%) | 1 (2.0%) | 2 (1.9%) | 1.0000 |
|  | Pain in extremity | 1 (1.9%) | 2 (4.0%) | 3 (2.9%) | 0.6104 |
| Neoplasm benign, malignant and unspecified | Neoplasms benign, mal, unspec - Oth spec | 4 (7.5%) | 3 (6.0%) | 7 (6.8%) | 1.0000 |
|  | Treatment related secondary malignancy | 1 (1.9%) | 0 (0.0%) | 1 (1.0%) | 1.0000 |
| Nervous system disorders | Dizziness | 0 (0.0%) | 1 (2.0%) | 1 (1.0%) | 0.4854 |
|  | Headache | 3 (5.7%) | 2 (4.0%) | 5 (4.9%) | 1.0000 |
|  | Neuralgia | 0 (0.0%) | 1 (2.0%) | 1 (1.0%) | 0.4854 |
|  | Nystagmus | 0 (0.0%) | 1 (2.0%) | 1 (1.0%) | 0.4854 |
|  | Paresthesia | 0 (0.0%) | 1 (2.0%) | 1 (1.0%) | 0.4854 |
|  | Peripheral motor neuropathy | 1 (1.9%) | 1 (2.0%) | 2 (1.9%) | 1.0000 |
|  | Seizure | 0 (0.0%) | 1 (2.0%) | 1 (1.0%) | 0.4854 |
|  | Spasticity | 1 (1.9%) | 0 (0.0%) | 1 (1.0%) | 1.0000 |
|  | Transient ischemic attacks | 0 (0.0%) | 1 (2.0%) | 1 (1.0%) | 0.4854 |
| Psychiatric disorders | Anxiety | 0 (0.0%) | 1 (2.0%) | 1 (1.0%) | 0.4854 |
|  | Depression | 2 (3.8%) | 1 (2.0%) | 3 (2.9%) | 1.0000 |
|  | Insomnia | 1 (1.9%) | 1 (2.0%) | 2 (1.9%) | 1.0000 |
| Renal and urinary disorders | Bladder spasm | 0 (0.0%) | 1 (2.0%) | 1 (1.0%) | 0.4854 |
|  | Renal and urinary disorders - Oth spec | 1 (1.9%) | 0 (0.0%) | 1 (1.0%) | 1.0000 |
|  | Renal calculi | 2 (3.8%) | 0 (0.0%) | 2 (1.9%) | 0.4955 |
|  | Urinary frequency | 1 (1.9%) | 1 (2.0%) | 2 (1.9%) | 1.0000 |
| Reproductive system and breast disorders | Dyspareunia | 1 (1.9%) | 0 (0.0%) | 1 (1.0%) | 1.0000 |
|  | Reproductive system and breast -Oth spec | 2 (3.8%) | 0 (0.0%) | 2 (1.9%) | 0.4955 |
|  | Vaginal hemorrhage | 1 (1.9%) | 0 (0.0%) | 1 (1.0%) | 1.0000 |
| Respiratory, thoracic, mediastinal disorders | Bronchospasm | 1 (1.9%) | 0 (0.0%) | 1 (1.0%) | 1.0000 |
|  | Cough | 1 (1.9%) | 1 (2.0%) | 2 (1.9%) | 1.0000 |
|  | Dyspnea | 1 (1.9%) | 1 (2.0%) | 2 (1.9%) | 1.0000 |
|  | Epistaxis | 1 (1.9%) | 1 (2.0%) | 2 (1.9%) | 1.0000 |
|  | Nasal congestion | 1 (1.9%) | 0 (0.0%) | 1 (1.0%) | 1.0000 |
|  | Pneumonitis | 0 (0.0%) | 1 (2.0%) | 1 (1.0%) | 0.4854 |
|  | Resp, thoracic, mediastinal - Oth spec | 1 (1.9%) | 1 (2.0%) | 2 (1.9%) | 1.0000 |
|  | Sinus disorder | 1 (1.9%) | 0 (0.0%) | 1 (1.0%) | 1.0000 |
|  | Sleep apnea | 1 (1.9%) | 0 (0.0%) | 1 (1.0%) | 1.0000 |
|  | Sneezing | 1 (1.9%) | 0 (0.0%) | 1 (1.0%) | 1.0000 |
|  | Sore throat | 1 (1.9%) | 2 (4.0%) | 3 (2.9%) | 0.6104 |
| Skin and subcutaneous tissue disorders | Erythema multiforme | 0 (0.0%) | 1 (2.0%) | 1 (1.0%) | 0.4854 |
|  | Rash maculo-papular | 3 (5.7%) | 1 (2.0%) | 4 (3.9%) | 0.6182 |
|  | Skin and subcut tissue disord - Oth spec | 7 (13.2%) | 1 (2.0%) | 8 (7.8%) | 0.0607 |
| Surgical and medical procedures | Surgical and medical proced - Oth spec | 2 (3.8%) | 1 (2.0%) | 3 (2.9%) | 1.0000 |
| Vascular disorders | Hot flashes | 1 (1.9%) | 0 (0.0%) | 1 (1.0%) | 1.0000 |
|  | Hypertension | 4 (7.5%) | 5 (10.0%) | 9 (8.7%) | 0.7367 |
|  | Thromboembolic event | 1 (1.9%) | 1 (2.0%) | 2 (1.9%) | 1.0000 |
| ^1^Fisher’s exact test | | | | | |

Table S4: Grade 2 Adverse Events

|  | | **Arm** | |  | |
| --- | --- | --- | --- | --- | --- |
| **MedDRA (Medical Dictionary for Regulatory Activities) SOC (system organ classes) (v12.0)** | **CTCAE (Common Terminology Criteria for Adverse Events)**  **Term (v4.0)** | MUC1 (N=53) | Placebo (N=50) | Total (N=103) | P-value^1^ |
| Cardiac disorders | Chest pain - cardiac | 2 (3.8%) | 1 (2.0%) | 3 (2.9%) | 1.0000 |
|  | Heart failure | 0 (0.0%) | 1 (2.0%) | 1 (1.0%) | 0.4854 |
|  | Myocardial infarction | 0 (0.0%) | 1 (2.0%) | 1 (1.0%) | 0.4854 |
| Ear and labyrinth disorders | Middle ear inflammation | 1 (1.9%) | 0 (0.0%) | 1 (1.0%) | 1.0000 |
|  | Tinnitus | 0 (0.0%) | 1 (2.0%) | 1 (1.0%) | 0.4854 |
|  | Vestibular disorder | 0 (0.0%) | 1 (2.0%) | 1 (1.0%) | 0.4854 |
| Eye disorders | Cataract | 0 (0.0%) | 1 (2.0%) | 1 (1.0%) | 0.4854 |
|  | Eye disorders - Other, specify | 1 (1.9%) | 0 (0.0%) | 1 (1.0%) | 1.0000 |
|  | Eyelid function disorder | 0 (0.0%) | 1 (2.0%) | 1 (1.0%) | 0.4854 |
| Gastrointestinal disorders | Abdominal pain | 2 (3.8%) | 0 (0.0%) | 2 (1.9%) | 0.4955 |
|  | Colitis | 1 (1.9%) | 0 (0.0%) | 1 (1.0%) | 1.0000 |
|  | Colonic obstruction | 1 (1.9%) | 0 (0.0%) | 1 (1.0%) | 1.0000 |
|  | Diarrhea | 2 (3.8%) | 1 (2.0%) | 3 (2.9%) | 1.0000 |
|  | Dyspepsia | 2 (3.8%) | 0 (0.0%) | 2 (1.9%) | 0.4955 |
|  | Gastric hemorrhage | 1 (1.9%) | 0 (0.0%) | 1 (1.0%) | 1.0000 |
|  | Gastroesophageal reflux disease | 0 (0.0%) | 1 (2.0%) | 1 (1.0%) | 0.4854 |
|  | Gastrointestinal disorders - Oth spec | 7 (13.2%) | 2 (4.0%) | 9 (8.7%) | 0.1618 |
|  | Hemorrhoids | 1 (1.9%) | 0 (0.0%) | 1 (1.0%) | 1.0000 |
|  | Nausea | 1 (1.9%) | 1 (2.0%) | 2 (1.9%) | 1.0000 |
|  | Pancreatitis | 1 (1.9%) | 0 (0.0%) | 1 (1.0%) | 1.0000 |
|  | Rectal pain | 1 (1.9%) | 0 (0.0%) | 1 (1.0%) | 1.0000 |
|  | Small intestinal obstruction | 1 (1.9%) | 0 (0.0%) | 1 (1.0%) | 1.0000 |
|  | Stomach pain | 1 (1.9%) | 0 (0.0%) | 1 (1.0%) | 1.0000 |
|  | Vomiting | 1 (1.9%) | 0 (0.0%) | 1 (1.0%) | 1.0000 |
| General disorders and administration site conditions | Fatigue | 1 (1.9%) | 0 (0.0%) | 1 (1.0%) | 1.0000 |
|  | Flu like symptoms | 1 (1.9%) | 0 (0.0%) | 1 (1.0%) | 1.0000 |
|  | Injection site reaction | 37 (69.8%) | 2 (4.0%) | 39 (37.9%) | **<.0001** |
|  | Non-cardiac chest pain | 0 (0.0%) | 1 (2.0%) | 1 (1.0%) | 0.4854 |
| Immune system disorders | Autoimmune disorder | 0 (0.0%) | 1 (2.0%) | 1 (1.0%) | 0.4854 |
| Infections and infestations | Bronchial infection | 1 (1.9%) | 1 (2.0%) | 2 (1.9%) | 1.0000 |
|  | Eye infection | 0 (0.0%) | 1 (2.0%) | 1 (1.0%) | 0.4854 |
|  | Infections and infestations - Oth spec | 2 (3.8%) | 2 (4.0%) | 4 (3.9%) | 1.0000 |
|  | Nail infection | 1 (1.9%) | 0 (0.0%) | 1 (1.0%) | 1.0000 |
|  | Otitis media | 0 (0.0%) | 1 (2.0%) | 1 (1.0%) | 0.4854 |
|  | Paronychia | 1 (1.9%) | 0 (0.0%) | 1 (1.0%) | 1.0000 |
|  | Sinusitis | 4 (7.5%) | 2 (4.0%) | 6 (5.8%) | 0.6789 |
|  | Skin infection | 1 (1.9%) | 0 (0.0%) | 1 (1.0%) | 1.0000 |
|  | Soft tissue infection | 1 (1.9%) | 0 (0.0%) | 1 (1.0%) | 1.0000 |
|  | Upper respiratory infection | 2 (3.8%) | 0 (0.0%) | 2 (1.9%) | 0.4955 |
|  | Urinary tract infection | 1 (1.9%) | 2 (4.0%) | 3 (2.9%) | 0.6104 |
| Injury, poisoning and procedural complications | Fall | 2 (3.8%) | 0 (0.0%) | 2 (1.9%) | 0.4955 |
|  | Fracture | 1 (1.9%) | 1 (2.0%) | 2 (1.9%) | 1.0000 |
|  | Inj, pois and proced complic - Oth spec | 2 (3.8%) | 0 (0.0%) | 2 (1.9%) | 0.4955 |
|  | Postoperative hemorrhage | 1 (1.9%) | 0 (0.0%) | 1 (1.0%) | 1.0000 |
|  | Wound complication | 1 (1.9%) | 0 (0.0%) | 1 (1.0%) | 1.0000 |
| Investigations | Alanine aminotransferase increased | 2 (3.8%) | 0 (0.0%) | 2 (1.9%) | 0.4955 |
|  | Investigations - Other, specify | 0 (0.0%) | 1 (2.0%) | 1 (1.0%) | 0.4854 |
|  | Lymphocyte count decreased | 1 (1.9%) | 0 (0.0%) | 1 (1.0%) | 1.0000 |
| Metabolism and nutrition disorders | Hyperglycemia | 1 (1.9%) | 0 (0.0%) | 1 (1.0%) | 1.0000 |
|  | Metabolism, nutrition disord - Oth spec | 1 (1.9%) | 0 (0.0%) | 1 (1.0%) | 1.0000 |
| Musculoskeletal and connective tissue disorders | Arthralgia | 4 (7.5%) | 2 (4.0%) | 6 (5.8%) | 0.6789 |
|  | Arthritis | 2 (3.8%) | 2 (4.0%) | 4 (3.9%) | 1.0000 |
|  | Back pain | 8 (15.1%) | 1 (2.0%) | 9 (8.7%) | **0.0318** |
|  | Joint range of motion decr cerv spine | 1 (1.9%) | 0 (0.0%) | 1 (1.0%) | 1.0000 |
|  | Musculoskeletal, conn tissue - Oth spec | 4 (7.5%) | 2 (4.0%) | 6 (5.8%) | 0.6789 |
|  | Myalgia | 0 (0.0%) | 1 (2.0%) | 1 (1.0%) | 0.4854 |
|  | Neck pain | 1 (1.9%) | 1 (2.0%) | 2 (1.9%) | 1.0000 |
|  | Pain in extremity | 0 (0.0%) | 1 (2.0%) | 1 (1.0%) | 0.4854 |
| Neoplasm benign, malignant and unspecified | Neoplasms benign, mal, unspec - Oth spec | 3 (5.7%) | 3 (6.0%) | 6 (5.8%) | 1.0000 |
|  | Treatment related secondary malignancy | 1 (1.9%) | 0 (0.0%) | 1 (1.0%) | 1.0000 |
| Nervous system disorders | Headache | 1 (1.9%) | 0 (0.0%) | 1 (1.0%) | 1.0000 |
|  | Neuralgia | 0 (0.0%) | 1 (2.0%) | 1 (1.0%) | 0.4854 |
|  | Nystagmus | 0 (0.0%) | 1 (2.0%) | 1 (1.0%) | 0.4854 |
|  | Paresthesia | 0 (0.0%) | 1 (2.0%) | 1 (1.0%) | 0.4854 |
|  | Peripheral motor neuropathy | 0 (0.0%) | 1 (2.0%) | 1 (1.0%) | 0.4854 |
|  | Seizure | 0 (0.0%) | 1 (2.0%) | 1 (1.0%) | 0.4854 |
|  | Transient ischemic attacks | 0 (0.0%) | 1 (2.0%) | 1 (1.0%) | 0.4854 |
| Psychiatric disorders | Depression | 1 (1.9%) | 1 (2.0%) | 2 (1.9%) | 1.0000 |
|  | Insomnia | 1 (1.9%) | 0 (0.0%) | 1 (1.0%) | 1.0000 |
| Renal and urinary disorders | Bladder spasm | 0 (0.0%) | 1 (2.0%) | 1 (1.0%) | 0.4854 |
|  | Renal and urinary disorders - Oth spec | 1 (1.9%) | 0 (0.0%) | 1 (1.0%) | 1.0000 |
|  | Renal calculi | 2 (3.8%) | 0 (0.0%) | 2 (1.9%) | 0.4955 |
|  | Urinary frequency | 0 (0.0%) | 1 (2.0%) | 1 (1.0%) | 0.4854 |
| Reproductive system and breast disorders | Reproductive system and breast -Oth spec | 1 (1.9%) | 0 (0.0%) | 1 (1.0%) | 1.0000 |
|  | Vaginal hemorrhage | 1 (1.9%) | 0 (0.0%) | 1 (1.0%) | 1.0000 |
| Respiratory, thoracic, mediastinal disorders | Bronchospasm | 1 (1.9%) | 0 (0.0%) | 1 (1.0%) | 1.0000 |
|  | Dyspnea | 0 (0.0%) | 1 (2.0%) | 1 (1.0%) | 0.4854 |
|  | Nasal congestion | 1 (1.9%) | 0 (0.0%) | 1 (1.0%) | 1.0000 |
|  | Resp, thoracic, mediastinal - Oth spec | 1 (1.9%) | 1 (2.0%) | 2 (1.9%) | 1.0000 |
|  | Sinus disorder | 1 (1.9%) | 0 (0.0%) | 1 (1.0%) | 1.0000 |
|  | Sleep apnea | 1 (1.9%) | 0 (0.0%) | 1 (1.0%) | 1.0000 |
|  | Sneezing | 1 (1.9%) | 0 (0.0%) | 1 (1.0%) | 1.0000 |
|  | Sore throat | 0 (0.0%) | 1 (2.0%) | 1 (1.0%) | 0.4854 |
| Skin and subcutaneous tissue disorders | Skin and subcut tissue disord - Oth spec | 6 (11.3%) | 1 (2.0%) | 7 (6.8%) | 0.1133 |
| Surgical and medical procedures | Surgical and medical proced - Oth spec | 2 (3.8%) | 1 (2.0%) | 3 (2.9%) | 1.0000 |
| Vascular disorders | Hot flashes | 1 (1.9%) | 0 (0.0%) | 1 (1.0%) | 1.0000 |
|  | Hypertension | 2 (3.8%) | 3 (6.0%) | 5 (4.9%) | 0.6722 |
|  | Thromboembolic event | 1 (1.9%) | 1 (2.0%) | 2 (1.9%) | 1.0000 |
| ^1^Fisher’s exact test |  |  |  |  |  |

Table S5: Grade 3 Adverse Events

|  | | **Arm** | |  | |
| --- | --- | --- | --- | --- | --- |
| **MedDRA (Medical Dictionary for Regulatory Activities) SOC (system organ classes) (v12.0)** | **CTCAE (Common Terminology Criteria for Adverse Events)**  **Term (v4.0)** | MUC1 (N=53) | Placebo (N=50) | Total (N=103) | P-value^1^ |
| Cardiac disorders | Chest pain - cardiac | 1 (1.9%) | 1 (2.0%) | 2 (1.9%) | 1.0000 |
|  | Heart failure | 0 (0.0%) | 1 (2.0%) | 1 (1.0%) | 0.4854 |
|  | Myocardial infarction | 0 (0.0%) | 1 (2.0%) | 1 (1.0%) | 0.4854 |
| Eye disorders | Cataract | 0 (0.0%) | 1 (2.0%) | 1 (1.0%) | 0.4854 |
| Gastrointestinal disorders | Abdominal pain | 1 (1.9%) | 0 (0.0%) | 1 (1.0%) | 1.0000 |
|  | Gastrointestinal disorders | 2 (3.8%) | 1 (2.0%) | 3 (2.9%) | 1.0000 |
|  | Pancreatitis | 1 (1.9%) | 0 (0.0%) | 1 (1.0%) | 1.0000 |
|  | Small intestinal obstruction | 1 (1.9%) | 0 (0.0%) | 1 (1.0%) | 1.0000 |
| Immune system disorders | Autoimmune disorder | 0 (0.0%) | 1 (2.0%) | 1 (1.0%) | 0.4854 |
| Injury, poisoning and procedural complications | Fall | 1 (1.9%) | 0 (0.0%) | 1 (1.0%) | 1.0000 |
|  | Postoperative hemorrhage | 1 (1.9%) | 0 (0.0%) | 1 (1.0%) | 1.0000 |
| Investigations | Investigations | 0 (0.0%) | 1 (2.0%) | 1 (1.0%) | 0.4854 |
| Musculoskeletal and connective tissue disorders | Arthralgia | 0 (0.0%) | 1 (2.0%) | 1 (1.0%) | 0.4854 |
|  | Arthritis | 1 (1.9%) | 1 (2.0%) | 2 (1.9%) | 1.0000 |
|  | Back pain | 0 (0.0%) | 1 (2.0%) | 1 (1.0%) | 0.4854 |
|  | Joint range of motion decreased cervical spine | 1 (1.9%) | 0 (0.0%) | 1 (1.0%) | 1.0000 |
|  | Musculoskeletal, connective tissue | 2 (3.8%) | 1 (2.0%) | 3 (2.9%) | 1.0000 |
| Neoplasm benign, malignant and unspecified | Neoplasms benign, mal, unspec | 1 (1.9%) | 2 (4.0%) | 3 (2.9%) | 0.6104 |
|  | Treatment related secondary malignancy | 1 (1.9%) | 0 (0.0%) | 1 (1.0%) | 1.0000 |
| Nervous system disorders | Peripheral motor neuropathy | 0 (0.0%) | 1 (2.0%) | 1 (1.0%) | 0.4854 |
| Renal and urinary disorders | Renal calculi | 1 (1.9%) | 0 (0.0%) | 1 (1.0%) | 1.0000 |
| Surgical and medical procedures | Surgical and medical procedures | 0 (0.0%) | 1 (2.0%) | 1 (1.0%) | 0.4854 |
| ^1^Fisher’s exact test | | | | | |
